# Investigation of bias due to selective inclusion of study effect estimates in meta-analyses of nutrition research

**DOI:** 10.1101/2022.11.01.22281823

**Authors:** Raju Kanukula, Joanne E McKenzie, Lisa Bero, Zhaoli Dai, Sally McDonald, Cynthia M Kroeger, Elizabeth Korevaar, Andrew Forbes, Matthew J Page

**Author notes:** Correspondence to: Dr. Matthew Page, Methods in Evidence Synthesis Unit, School of Public Health and Preventive Medicine, Monash University, 553 St Kilda Road, Melbourne, Victoria, 3004, Australia.

## Abstract

We aimed to explore, in a sample of systematic reviews with meta-analyses of the association between food/diet and health-related outcomes, whether systematic reviewers selectively included study effect estimates in meta-analyses when multiple effect estimates were available. We randomly selected systematic reviews of food/diet and health-related outcomes published between January 2018 and June 2019. We selected the first presented meta-analysis in each review (index meta-analysis), and extracted from study reports all study effect estimates that were eligible for inclusion in the meta-analysis. We calculated the Potential Bias Index (PBI) to quantify and test for evidence of selective inclusion. The PBI ranges from 0 to 1; values above or below 0.5 suggest selective inclusion of effect estimates more or less favourable to the intervention, respectively. We also compared the index meta-analytic estimate to the median of a randomly constructed distribution of meta-analytic estimates (i.e. the estimate expected when there is no selective inclusion). Thirty-nine systematic reviews with 312 studies were included. The estimated PBI was 0.49 (95% CI 0.42 to 0.55), suggesting that the selection of study effect estimates from those reported was consistent with a process of random selection. In addition, the index meta-analytic effect estimates were similar, on average, to what we would expect to see in meta-analyses generated when there was no selective inclusion. Despite this, we recommend that systematic reviewers report the methods used to select effect estimates to include in meta-analyses, which can help readers understand the risk of selective inclusion bias in the systematic reviews.

## 1. BACKGROUND

Systematic reviews (SRs) synthesise the findings of studies that address a specific research question. By attempting to include all available evidence meeting pre-specified eligibility criteria, SRs can provide more valid evidence for healthcare decision making than non-systematic literature reviews or ad hoc findings of single studies^1^. It is now commonplace for SRs to underpin clinical practice guidelines and policy decisions^2^. Furthermore, SRs play a critical role in identifying research gaps and directing future research. However, the validity of the findings from SRs is, in part, dependent on the use of methods that minimise bias in the SR process.

Systematic reviewers commonly encounter multiple effect estimates from a primary study that are eligible for inclusion in a particular meta-analysis (which we refer to as ‘multiplicity of results within studies’)^3–5^. For example, in a particular primary study, mean differences between intervention groups for three different quality of life questionnaires measured at 1-, 2- and 3-months follow-up may be reported, all of which are eligible for a planned meta-analysis of quality of life at 0-3 months follow-up. Naïve inclusion of all nine estimates in the meta-analysis introduces statistical dependency in the meta-analytic dataset, which can lead to misleading results^6^.

Two general approaches for dealing with multiplicity of study results in a meta-analysis include the integrative and reductionist approach^6^. Integrative approaches aim to include multiple effect estimates per study in the meta-analysis, using statistical methods such as multilevel modelling^7^ and robust variance estimation^8^. These approaches are not the focus of this paper. Reductionist approaches, which are the focus of this paper, involve reducing the data so that only one effect estimate per study is included in a particular meta-analysis. Best practice for reductionist approaches is to pre-specify “eligibility criteria”, indicating which results are eligible for inclusion in the meta-analysis, along with “decision rules”, that specify the methods used to select the result for inclusion, when multiple are available^5, 9^. These rules articulate a preference for one estimate over another, and typically this will be based on perceived credibility of the estimate (e.g. selecting based on the most valid and reliable scales), or for clinical (e.g. selecting clinically relevant measures) or methodological (e.g. selecting the most consistently used measure) reasons. In practice, however, such rules are not always specified in advance^9^, leading to the potential for selection of study effect estimates based on the observed result itself (e.g. selecting the largest estimate, or the one with the smallest P-value). This is known as ‘selective inclusion of results’^4, 9^, and has to potential to bias meta-analysis results.

To our knowledge, only one study has investigated bias due to selective inclusion of results in meta-analyses^10^. This study focused on SRs (published between 2010 and 2012) of randomised trials examining the effects of interventions for arthritis or depressive or anxiety disorders. The authors constrained their investigation to the first presented meta-analysis of a continuous outcome in each review. They concluded that in this random sample of SRs, there was no clear evidence of selective inclusion of effect estimates, and that any potential selective inclusion did not have a meaningful impact on the meta-analytic effects. However, it is unclear whether these findings apply to other clinical conditions, outcome types (e.g. binary), study designs, and more recently published reviews.

Systematic reviews of nutrition research provide an opportunity to extend the previous investigation of bias due to selective inclusion, because these reviews often include study designs beyond randomised trials, different outcome types, and include results from multiple analyses that attempt to adjust for potential confounding variables (in the case of non-randomised studies). Furthermore, given the importance of SRs of nutrition research in informing recommendations in dietary guidelines and public health policy, assessment of whether there is evidence of selective inclusion bias is required. Therefore, the objectives of this study were to explore, in a sample of SRs with meta-analyses of the association between food/diet and health-related outcomes: (i) whether systematic reviewers selectively included study effect estimates in meta-analyses when multiple effect estimates were available, and; (ii) what impact selective inclusion of study effect estimates may have on meta-analytic effects.

## 2. METHODS

The study protocol for the ROBUST study has been published^11^. The ROBUST study, in addition to objectives (i) and (ii) above, aims to investigate the risk of bias due to missing results (objective (iii) in the study protocol). The results of this latter investigation will be reported in a subsequent manuscript. An investigation of the extent of multiplicity of results in the primary studies of the sample of SRs included in the ROBUST study, has been published^5^. Here, we provide an overview of the methods relevant for our investigation of bias due to selective inclusion of results, with deviations from the planned methods presented in Supplementary Table S1.

### 2.1. Eligibility criteria, search and selection of SRs

We included reviews that satisfied the definition of an SR, according to the 2019 edition of the Cochrane Handbook for Systematic Reviews of Interventions^12^, that had explicitly stated methods of study identification (e.g. a search strategy) and of study selection (e.g. eligibility criteria and selection process), and included a meta-analysis. We included such SRs with a meta-analysis that:

- included studies that enrolled, regardless of their age and background, (a) people who were generally healthy, or (b) a mixture of generally healthy people and people with diet-related risk factors (e.g. overweight, high blood pressure) or people with a particular health condition (e.g. type II diabetes or cardiovascular disease), or (c) people with non-specified health status;
- included randomised trials or non-randomised studies that assessed the effects of at least one type of food (e.g. nuts, red meat) or at least one dietary pattern (e.g. vegan) on any continuous (e.g. weight) or non-continuous (e.g. cardiovascular disease incidence) health-related outcome;
- were published in English between 1 January 2018 and 30 June 2019 (i.e. within 18 months prior to the drafting of our study protocol);
- provided citations for all included studies in the SR, and;
- presented the summary statistics or an effect estimate and its precision (e.g. standard error or 95% confidence interval) for each study included in a meta-analysis, and the meta-analytic summary effect estimate and its precision in the text or forest plot.

The exclusion criteria can be found in our published protocol^11^.

We searched for eligible SRs indexed in the PubMed and Epistemonikos databases from 1 January 2018 to 30 June 2019 (search strategies reported in supplementary Table S2), and exported unique records to Microsoft Excel. Rather than screening all records, we screened, in random order, as many records as we needed to reach our target sample of SRs. Following piloting of the eligibility criteria, two investigators (MJP and one of CMK, ZD or SM) independently screened titles and abstracts (in batches of 500 records and in random order) and their potentially eligible full-text reports. This screening process was repeated until we identified 50 eligible SRs, including 25 meta-analyses of continuous and 25 meta-analyses of non-continuous outcomes. If the total number of eligible SRs exceeded this target at the end of a batch, we planned to sample 25 SRs of each type randomly. Our target of 50 SRs was primarily selected for feasibility reasons given our available resources to conduct all components of the ROBUST study, which was informed by the time taken to conduct a previous similar study^10^. Any discrepancies in screening decisions at each stage were resolved via discussion between investigators or by consultation with another investigator (JM) where necessary.

One investigator (MJP) selected from each included SR one pairwise meta-analysis (i.e. comparing one intervention/exposure against another) of aggregate data for inclusion in the present study. The selected meta-analysis was the first meta-analytic result mentioned in the review (regardless of its placement in the manuscript). Henceforth, we refer to the selected meta-analysis as the “index meta-analysis”. Initially, the index meta-analysis was selected irrespective of the outcome domain (e.g. quality of life, prostate cancer), effect measure (e.g. risk ratio, mean difference), meta-analytic model (fixed-effect, random-effects) or the number and type of included studies (i.e. randomised or non-randomised study). However, following the selection of 50 index meta-analyses, we determined that they included 553 studies (range 2-55 studies per meta-analysis), which is more than twice what we had anticipated based on our previous study^10^. For feasibility reasons, we therefore chose to restrict inclusion to only those SRs with an index meta-analysis including fewer than 20 studies. For each included index meta-analysis, we retrieved the reports of all included studies (see the protocol for further details^11^).

### 2.2 Data collection and management

A data collection form was developed in REDCap^13^ (see Supplementary Table S3). Following piloting, two investigators (RK and one of ZD, SM, CMK, EK, LB and MJP) independently collected data from a random sample of half of the index meta-analyses and their included studies. For the remaining index meta-analyses and their included studies, data were collected by one investigator (RK) and verified for accuracy by another investigator (MJP). Any discrepancies were resolved through discussions between the two investigators or through adjudication by a third investigator (JM) if necessary.

An overview of the data items and the sources these were obtained from (i.e. systematic review protocol, systematic review or study report) is presented in Supplementary Table S4; further details are available in the protocol^11^. For data extracted from the reports of studies included in the index meta-analysis, we extracted all outcome data eligible for inclusion in the index meta-analysis. This was determined by the eligibility criteria and decision rules stated in the SR protocol when available. For example, if the systematic reviewers pre-defined in the SR protocol that for the meta-analysis of weight gain, they would only compare the “highest versus lowest intake of fish”, and would consider only data at 8 weeks follow-up when data were available at multiple time points, we only extracted data for that comparison and time point, regardless of whether data for other time points and other comparisons for the same outcome were available in the study reports. If there was no SR protocol available, or if there were no eligibility criteria or decision rules to select results specified in the SR protocol, we extracted all study outcome data based on the comparison and outcome specified in the SR. For example, if the meta-analysis in the SR was described as “fish intake versus none on weight at 6 months”, we extracted all data on weight at 6 months for comparisons of any level of intake of fish versus none, results that were unadjusted and adjusted (for potential confounding variables), and results from all analysis samples (e.g. per-protocol, intention-to-treat). We ignored any eligibility criteria or decision rules to select results that appeared in the SR, as we could not determine whether they were pre-specified. Additional rules for determining which data to collect are outlined in the protocol^11^.

### 2.3 Data analysis

#### 2.3.1 Descriptive analysis

We summarised the characteristics of SR protocols, SRs, index meta-analyses and included studies using descriptive statistics. For categorical variables, we present frequencies and percentages. For continuous variables, we report medians with interquartile ranges (IQRs).

#### 2.3.2. Quantification and testing for evidence of selective inclusion of results

We used the ‘Potential Bias Index’ (PBI) to quantify and test for evidence of selective inclusion^14^. Mathematical details of the construction of the PBI and associated statistical test are provided in Supplementary Table S5 and a worked example in Supplementary Table S6. In brief, this index is based on ordering the effect estimates in each primary study according to their magnitude and direction of effect, and then determining the position within that order where the effect estimate included in the index meta-analysis sits. A rank of 1 is assigned to the smallest effect estimate and a rank equal to the number of effect estimates is assigned to the largest effect estimate. Since the number of effect estimates varies across studies, we rescale the ranks of the selected effect estimates to reflect their relative positioning (in ranking units from 0 to 1) between the smallest and largest effect estimates^14^. The PBI is then calculated as the weighted average rank position of the selected effect estimates across all studies (that have at least two effect estimates) included in the index meta-analyses, with the weights being the number of effect estimates available per study. This weighting system, therefore, attributes greater priority to the rank positions of effect estimates where there are a larger number of effect estimates to choose from.

The PBI ranges from 0 to 1. The PBI has the value 1 when the effect estimate that is most favourable to the experimental intervention/exposure (of those reported) is always selected for inclusion from each study. By “most favourable” we mean the effect estimate that suggested the most benefit or least harm of the intervention/exposure. Conversely, the PBI would have a value of 0 when the effect estimate that is least favourable to the experimental intervention/exposure (of those reported) is always selected for inclusion from each study. Several methods for selecting effect estimates, from those reported, are acceptable in terms of not introducing bias through the selection process. These include when systematic reviewers (i) randomly select one of the reported effect estimates, (ii) select one of the reported effect estimates based on some clinical or methodological rationale or (iii) select the median of the reported effect estimates. Selection methods (ii) and (iii) are akin to selection method (i) because we expect the distribution of selected effect estimates would be consistent with what we would observe under purely random selection, and so on average, the selected effect estimates would be at the middle rank position, and the PBI would take the value of 0.5. A PBI of 0.5, therefore, suggests that there is no selective inclusion of the most or least favourable reported effect estimates.

For meta-analyses comparing different levels or patterns of intake of the same food (e.g. red meat consumed 5 days per week versus red meat consumed once a week), we determined from the text of the review whether the systematic reviewers hypothesised that the higher or lower category would have the most benefit or least harm, and ranked study effect estimates based on their favourability to the category of consumption considered to be most beneficial/least harmful. For meta-analyses comparing different foods/diets (e.g. vegan versus vegetarian diet), we determined from the text of the review which intervention/exposure the systematic reviewers were most interested in evaluating (which we considered the experimental intervention/exposure), and ranked the study effect estimates based on their favourability to the experimental intervention/exposure. Given there was more uncertainty in determining the experimental intervention/exposure in reviews comparing different food/diets, we performed an *a-priori* specified sensitivity analysis where we excluded meta-analyses comparing different foods/diets to examine the impact on the PBI. One investigator (RK) assigned a ranking to each effect estimate extracted from each study included in the index meta-analyses, and these rankings were verified by another investigator (MJP).

We ran a statistical test based on the PBI that has been constructed to test whether the observed selection of effect estimates was consistent with randomness of selection^14^. Confidence intervals (95%) for the PBI were obtained by bootstrap resampling^15^. Index meta-analyses that included no studies with multiplicity of effect estimates were excluded from all PBI analyses, given there is no potential for selective inclusion of results in such meta-analyses.

We conducted subgroup analyses to explore whether the availability of an SR protocol or register entry, SRs funded by food industry, outcome types for the index meta-analysis, or SRs having at least one author disclosing a financial conflict of interest of any type, modified the PBI. The confidence intervals and p value for the difference in PBI between subgroups were constructed using bootstrap methods^15^. A similar approach was used to examine whether the PBI was modified by the number of available effect estimates, where a regression of PBI on the number of available effect estimates was fitted (for details, see the study protocol^11^).

We undertook a series of pre-specified sensitivity analyses to investigate whether the PBI was robust to certain assumptions. For SRs without protocols or register entries, our primary calculation of the PBI was based on the set of study effect estimates that were compatible with the assumption of ‘no pre-specified eligibility criteria or decision rules’. However, we also performed a sensitivity analysis where study effect estimates that were compatible only with the eligibility criteria and decision rules in the methods sections of the SR were included, to examine if this restriction affected the PBI.

We anticipated that in some study reports, only an effect estimate and its standard error or 95% confidence interval would be presented (that is, the number of events or means and standard deviations per group would not be available). In this circumstance, algebraic manipulation was required to include the result in a meta-analysis. Algebraic manipulation may be considered challenging by some systematic reviewers, so effect estimates requiring algebraic manipulation may not have been considered by reviewers in the set of effect estimates to potentially include in the meta-analysis. For the primary calculation of the PBI, we excluded study effect estimates that required algebraic manipulation; however, we performed a sensitivity analysis to explore whether the PBI was modified when we included these study effect estimates.

At the suggestion of a peer-reviewer, we undertook a further sensitivity analysis to examine whether the PBI estimates varied across the meta-analyses, as might arise if some systematic reviewers selectively included effect estimates and others did not. This involved calculating an estimate of the PBI per meta-analysis (using the formula for the PBI and its variance in Supplementary Table S5), and meta-analysing these estimates using a random-effects meta-analysis model. The between-study variance was estimated using the restricted maximum likelihood (REML) estimator. The Hartung-Knapp-Sidik-Jonkman (HKSJ) confidence interval method was used to calculate uncertainty in the estimated average PBI^16, 17^. We quantified statistical inconsistency using the *I*^2^ statistic^18^, and calculated a prediction interval, which provides a summary of the spread of underlying PBIs in the meta-analyses included in our analysis^19^.

We conducted a fixed-effect meta-analysis of the PBI obtained in the current study with that estimated in the previous study by Page et al.^10^. We synthesised estimates of the PBI using a fixed-effect meta-analysis model because the number of included studies (n=2) was too small to adequately estimate the between-study variance.

#### 2.3.3. Investigation of the impact of selection of study effect estimates on meta-analysis effects

We investigated the impact of selection of study effect estimates on the magnitude of the resulting meta-analytic effect estimates. For continuous outcomes, we expressed all study effect estimates as standardised mean differences (SMDs). For non-continuous outcomes, we expressed all study effect estimates as odds ratios (ORs). When study authors reported risk ratios or hazard ratios for non-continuous outcomes, we extracted these estimates and made the assumption that these would provide a reasonable approximation to an odds ratio^20^, given the outcomes were rare in the included reviews. We standardised the direction of effects so that ORs < 1 or SMDs < 0 represented effects that are more favourable to the experimental intervention/exposure.

For each of the meta-analyses of continuous outcomes, we calculated all possible meta-analytic SMDs from all combinations of available study effect estimates. All possible meta-analytic SMDs were generated using a random-effects meta-analysis model, with the between-study variability estimated using the REML estimator. When the number of possible combinations was prohibitively large to calculate all combinations (i.e. >30,000), we generated a random sampling distribution of 5,000 meta-analytic SMDs. This was achieved by randomly selecting (with equal probability) an effect estimate for inclusion from each study comparison within a meta-analysis, meta-analysing the chosen effects to yield one meta-analytic result, and repeating this process 5,000 times. For each distribution of generated meta-analyses, we calculated (i) the median of all possible meta-analytic SMDs, which represents the median of a distribution where study effect estimates were not selectively included, and (ii) the difference between the index meta-analytic SMD and the median meta-analytic SMD.

We used non-parametric statistics to describe the differences between the index and median meta-analytic SMDs. We also synthesised these differences using a random-effects meta-analysis model, with the meta-analytic weights based on the variance of the index meta-analytic SMD estimate and the between-study variability estimated using the REML estimator. The HKSJ confidence interval method was used to calculate uncertainty in the combined differences^16, 17^, and we quantified statistical inconsistency using the I^2^ statistic^18^. When the difference between the index and median meta-analytic SMD was minimal, we concluded that on average, the index meta-analytic effect estimates were similar to what we would expect to see in meta-analyses generated when there was no selective inclusion (a worked example is provided in Supplementary Table S7). We repeated the above analyses for each of the meta-analyses of non-continuous outcomes (i.e. binary, count, time-to-event) by calculating meta-analytic ORs. Finally, we undertook an analysis that included all meta-analyses (i.e. including meta-analyses of all outcome types) by first converting log ORs to SMDs by dividing the log ORs by π/√3 (= 1.814)^21^. For this analysis, we also calculated all possible meta-analytic estimates using a fixed-effect model as a sensitivity analysis to examine whether the meta-analysed difference and its confidence interval were affected by the meta-analysis model. We also performed a sensitivity analysis where the meta-analyses of risk ratios and hazard ratios were excluded because of the assumption that such estimates provide a good approximation to the OR. All analyses were undertaken using Stata version 16^22^.

## 3. RESULTS

Our search yielded a total of 7,167 references from PubMed and Epistemonikos. After removing 908 duplicates, the remaining 6259 references were randomly sorted, and we screened the titles and abstracts of the first 3,013 references. Of these, 2,777 were excluded for not meeting the eligibility criteria, leaving 236 for full-text screening. Of these, 99 SRs met our eligibility criteria, including 25 SRs with a meta-analysis of a continuous outcome and 74 with a meta-analysis of a non-continuous outcome. Initially, all SRs with a continuous outcome were included, and 25 of the 74 SRs with a non-continuous outcome were randomly selected. However, from these 50 SRs, eight were excluded (6 continuous, 2 non-continuous), because the index meta-analysis had 20 or more studies, leaving 42 included SRs^23–64^. After extracting data from the included studies of these 42 SRs, we excluded a further three SRs because we were unable to identify the relevant outcome data in any of the included primary study reports^62^, or none of the included studies had multiplicity of effect estimates^63 64^. This left us with 39 SRs included in the analysis of selective inclusion (Figure 1).

**Figure 1.**
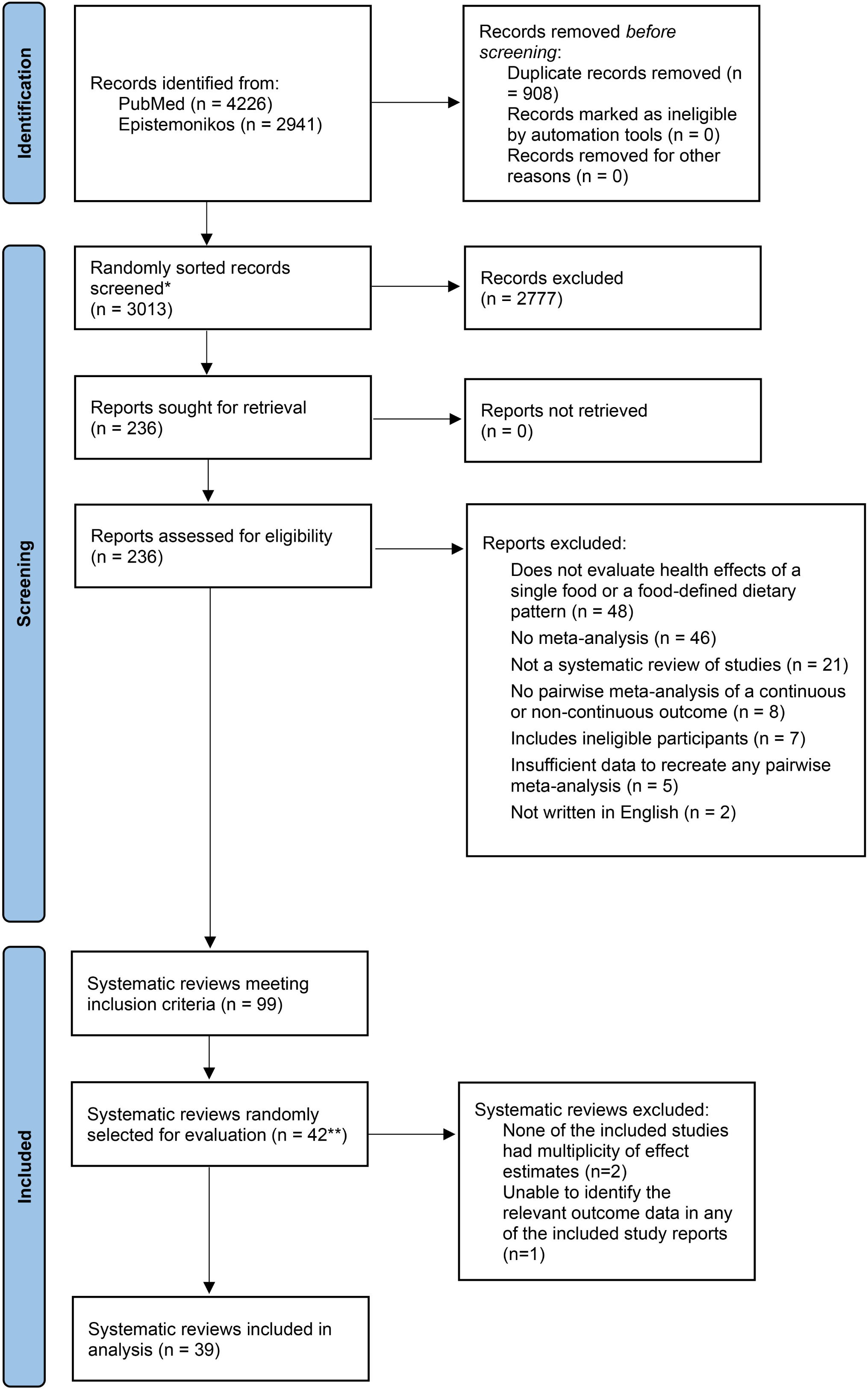
PRISMA 2020 flow diagram of identification, screening and inclusion of systematic reviews. *Of the 6259 unique titles and abstracts, we only needed to screen 3,013 randomly sorted titles and abstracts to reach our target sample size. **We initially drew a random sample of 50 systematic reviews, but post-hoc excluded eight systematic reviews with 20 or more included studies in the index meta-analysis to reduce workload.

### 3.1 Characteristics of included meta-analyses

Of the 39 SRs, two had published protocols, and 11 were registered in PROSPERO (Table 1). Most index meta-analyses included only non-randomised studies (64%, 25/39), 31% (12/39) included only randomised trials, and the remaining two (5%) included studies of both designs (Table 1). Only two reviews (5%) were funded by food industry and authors of six (15%) reviews had a financial conflict of interest. The types of included study designs differed by outcome type. In the 17 meta-analyses of a continuous outcome, 12 included randomised trials only, three included non-randomised studies only, and two included both designs. Of the 22 meta-analyses of a non-continuous outcome, all included non-randomised studies only. Nearly all index meta-analyses were fitted using a random-effects model (92%, 36/39). The 39 index meta-analyses included a total of 312 studies with 386 comparisons (since some studies contributed multiple comparisons to the same meta-analysis when, for example, multiple foods or diets were compared), with a median of seven studies (IQR 5-11; range 2-17) per meta-analysis.

**Table 1:**
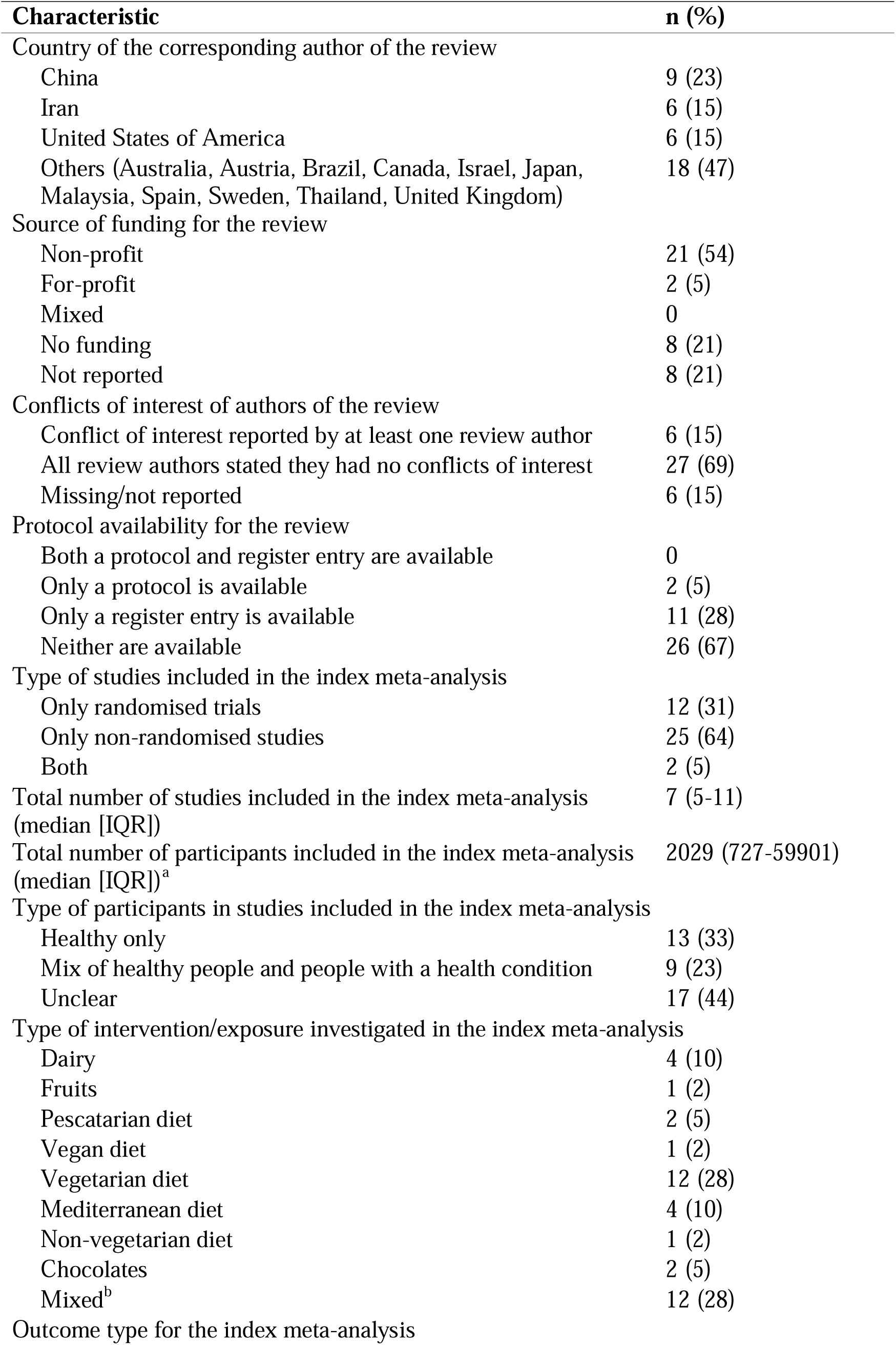
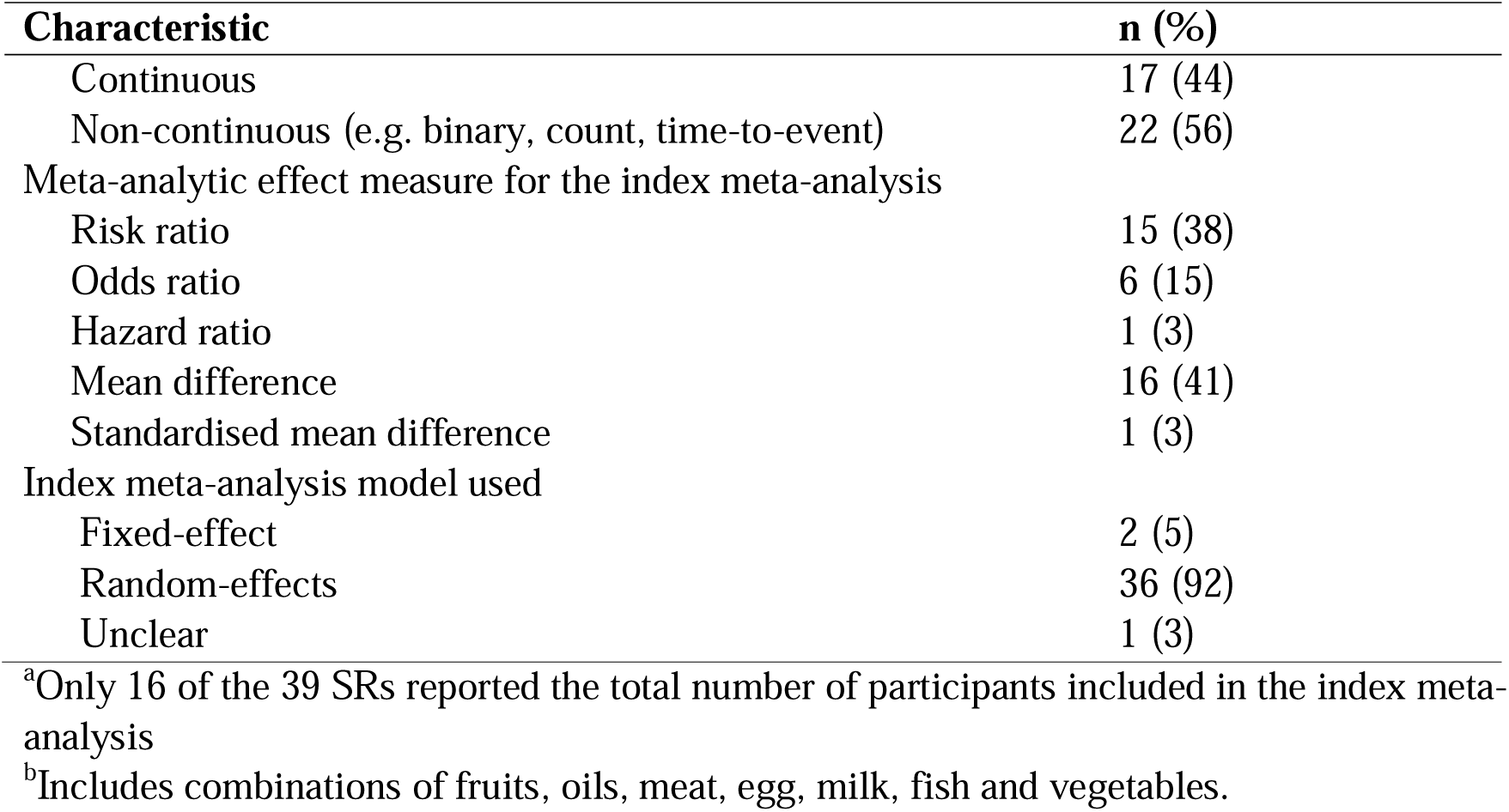
Characteristics of the systematic reviews and index meta-analyses (N=39)

### 3.2 Multiplicity of effect estimates in study reports and methods to select effect estimates

Of the 386 comparisons, 224 (58%) had multiple effect estimates that were eligible for inclusion in a particular meta-analysis. In these 224 comparisons, there was a median of 2 (IQR 2-4; range 2-21) eligible effect estimates per comparison. Descriptive statistics at the meta-analysis level reflected those at the study level; per meta-analysis, a median of 63% of comparisons had multiplicity (IQR 50-83%; range 6-100%). The most common types of multiplicity arose from the availability of unadjusted and one or more covariate-adjusted analyses (39% of studies) and multiple intervention/exposure groups (22% of studies) (Table 2).

**Table 2:** Number (%) of studies (N=312) with different types of multiplicity of results.

| Type of multiplicity | n (%)<sup>a</sup> |
| --- | --- |
| Any | 214 (69) |
| Multiple measurement instruments for an outcome domain | 0 |
| Multiple intervention/exposure groups | 68 (22) |
| Multiple time points | 14 (4) |
| Both final and change from baseline values | 27 (9) |
| Analyses undertaken on multiple samples (e.g., ITT and per-protocol) | 4 (1) |
| Unadjusted and one or more covariate-adjusted analyses | 121 (39) |
| Both period and paired analyses in crossover randomized trials | 0 |
| Multiple definitions of an event | 4 (1) |
| Multiple subgroups | 24 (8) |
| Other | 12 (4) |

<sup>a</sup>Percentages do not sum to 100 because some studies had multiple sources of multiplicity of effect estimates. The denominator was the number of studies, not study comparisons. Therefore, each multi-arm study that contributed multiple comparisons to a meta-analysis (e.g. 'high versus low intake of milk' and 'high versus low intake of cheese' included in a meta-analysis of the effect of dairy intake on risk of prostate cancer) was counted only once.

The types of methods to select effect estimates to include in meta-analyses that were documented in the SR protocols and SRs varied considerably (Table 3 and Supplementary Table S8). For example, in 1/13 protocols a decision rule for any type of analysis was provided, while such a rule appeared in 14/39 in reviews. In all cases where a decision rule was reported, the rule articulated selection of a *specific* eligible effect estimate (Table 3). In no cases did review authors articulate *non-specific* selection of an effect estimate (e.g. through random selection, or calculating an average of the eligible effect estimates).

**Table 3:** Content of decision rules reported in systematic review protocols and systematic reviews.

| Type of decision rule | SR protocols | SRs |
| --- | --- | --- |
|  | n (%)<br>N=13 | n (%)<br>N=39 |
| Measurement of outcome |  |  |
| Not applicable as the index outcome was defined by a particular instrument | 1 (8) | 1 (3) |
| No decision rule | 12 (92) | 38 (97) |
| Intervention/exposure and control groups |  |  |
| If the same intervention/exposure, but more than one intake level, then select the highest/lowest level | 0 | 17 (44) |
| If the same intervention/exposure, but with two intake levels, then combine into one group | 1 (8) | 2 (5) |
| If two different interventions/exposures, include each as separate comparison | 1 (7) | 2 (5) |
| Other<sup>a</sup> | 0 | 1 (3) |
| No decision rule | 11 (84) | 17 (44) |
| Time points |  |  |
| End of the study | 0 | 1 (3) |
| Other specific time point | 0 | 1 (3) |
| No decision rule | 13 (100) | 37 (95) |
| Final versus change from baseline values |  |  |
| Final values preferred | 0 | 0 |
| Change from baseline values preferred | 0 | 5 (13) |
| No decision rule | 13 (100) | 34 (87) |
| Unadjusted versus covariate adjusted analyses |  |  |
| Unadjusted analyses preferred to adjusted | 0 | 1 (3) |
| Adjusted analyses preferred to unadjusted | 0 | 3 (8) |
| Most adjusted analyses preferred if multiple adjusted analyses were reported | 0 | 2 (5) |
| No decision rule | 13 (100) | 33 (85) |
| Period or paired analyses in cross-over trials |  |  |
| First period results only | 0 | 1 (3) |
| No decision rule | 13 (100) | 38 (97) |
| Analyses undertaken on multiple samples |  |  |
| Intention-to-treat (ITT) analysis preferred | 0 | 0 |
| If there was missing data, modified ITT analysis preferred over per-protocol or as-treated analysis | 0 | 0 |
| Per protocol analysis preferred | 0 | 0 |
| As-treated analysis preferred | 0 | 0 |
| No decision rule | 13 (100) | 39 (100) |

<sup>a</sup>Classified based on the authors' description (e.g. when two or more healthy dietary patterns were presented, researchers generally considered a healthy pattern one that mostly included protective foods, such as vegetables, fruits, fish, legumes, whole grains, and low-fat dairy products)

### 3.3 Evidence of selective inclusion of study effect estimates

The estimated PBI was 0.49 (95% CI 0.42 to 0.55; two tailed p-value of 0.64) (Table 4). This suggests that the selection of study effect estimates from those reported was consistent with a process of random selection. The PBI for SRs *with* a protocol/register entry was 0.16 units higher (95% CI 0.02 to 0.29) than the PBI for SRs *without* a protocol/register entry, suggesting that selective inclusion of the most favourable effect estimates was more likely to occur in SRs with a protocol/register entry. The PBI was not modified by the outcome type of the index meta-analysis, funding of the SR or financial conflicts of interest of SR authors (Table 2), and was robust to the pre-specified sensitivity analyses (Supplementary Table S9). Furthermore, the peer-reviewer-suggested sensitivity analysis examining whether the PBI estimates varied across the meta-analysis, provided no evidence that the PBI estimates varied (I^2^ = 0%; f^. = 0), and yielded the same combined PBI estimate (Supplementary Table S10, Supplementary Figure S10).

**Table 4.** Primary and subgroup analyses for the PBI results.

| **PBI analyses** | **Number of comparisons** | **Number of meta-analyses** | **PBI (95% CI<sup>a</sup>)** |
| --- | --- | --- | --- |
| Primary analysis<sup>b</sup> | 206 | 38 | 0.49 (0.42-0.55) |
| Availability of a SR protocol/register entry |  |  |  |
| With a SR protocol/register entry | 40 | 13 | 0.61 (0.49-0.73) |
| Without a SR protocol/register entry | 166 | 25 | 0.46 (0.38-0.53) |
| Subgroup difference<sup>c</sup> = 0.16 (95% CI 0.02 to 0.29; two-tailed p value of test of interaction 0.03) |  |  |  |
| Outcome type for index meta-analysis |  |  |  |
| Continuous | 45 | 16 | 0.57 (0.45-0.68) |
| Non-continuous | 161 | 22 | 0.47 (0.40-0.54) |
| Subgroup difference<sup>c</sup> = 0.10 (95% CI -0.04 to 0.24; two-tailed p value of test of interaction 0.15) |  |  |  |
| Funding type for SR |  |  |  |
| Non-industry | 196 | 36 | 0.48 (0.42-0.54) |
| Industry | 10 | 2 | 0.51 (0.25-0.77) |
| Subgroup difference<sup>c</sup> = -0.03 (95% CI -0.27 to 0.26; two-tailed p value of test of interaction 0.84) |  |  |  |
| Financial conflict of interest of SR authors |  |  |  |
| No SR author had a conflict of interest | 166 | 32 | 0.48 (0.42-0.55) |
| ≥SR author had a conflict of interest | 40 | 6 | 0.49 (0.36-0.63) |
| Subgroup difference<sup>†</sup> = -0.01 (95% CI -0.16 to 0.14; two-tailed p value of test of interaction 0.87) |  |  |  |

<sup>a</sup>Percentile-based CIs for the PBI were constructed using bootstrap methods by resampling individual studies 2000 times.
<sup>b</sup>There are 206 (rather than 224) comparisons in the primary analysis, because the primary analysis omits comparisons for which inclusion of effect estimates required algebraic manipulation.
<sup>c</sup>The confidence limits and p value for the difference in PBI between subgroups was constructed using bootstrap methods from 2000 replicates.

The linear regression exploring the relationship between the number of available effects estimates and the PBI suggested that for every unit increase in the number of effect estimates, the PBI was predicted to reduce by −0.014 (95% CI −0.018 to −0.010; see Supplementary Figure S1). For example, the predicted PBI was 0.50 (95% CI 0.42 to 0.59) when there were two available effect estimates, and 0.41 (95% CI 0.25 to 0.57) when there were four effect estimates.

Meta-analysis of the PBI estimate of the previous study by Page et al.^10^ and our study resulted in a combined PBI of 0.53 (95% CI 0.48 to 0.58) (Supplementary Figure S2).

### 3.4 Impact of selection of study effect estimates on meta-analytic SMDs

The median number of possible meta-analytic SMDs arising from meta-analysing all combinations of study effect estimates was 288 (across the 39 meta-analyses); however, there was wide variation in this number (IQR 12-2880, range 2–20155392; Table 5). For most meta-analyses, the range of possible meta-analytic SMDs which could be calculated (each fitted using a random-effects model) was narrow (Table 5, column 5). The median difference between the largest and smallest possible meta-analytic SMD was 0.09 SD units (IQR 0.04 to 0.15; range 0.01 to 1.37; Table 5 and Figure 2).

**Figure 2.**
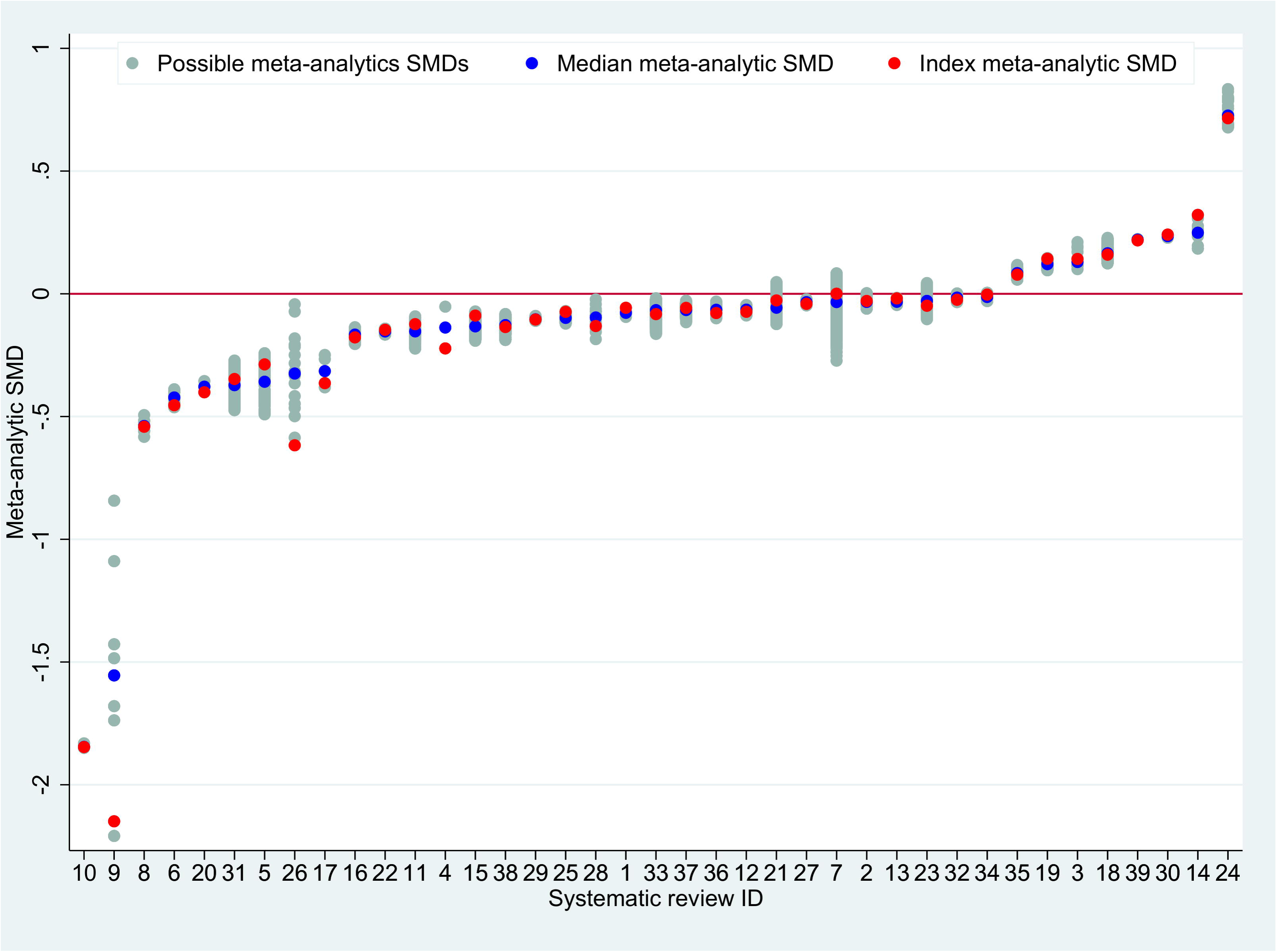
Range of possible meta-analytic standardised mean differences (SMDs) per index meta-analysis, with median and index meta-analytic SMD.

**Table 5:** Number (%) of studies with multiplicity, number of possible meta-analytic standardised mean differences (SMDs), and differences between meta-analytic SMDs for each index meta-analysis (N=39)

| **SR ID** | **Number of study comparisons** | **Number (%) of study comparisons with multiplicity** | **Number of meta-analytic SMDs** | **Largest minus smallest meta-analytic SMD** | **Index minus median meta-analytic SMD<sup>a</sup>** |
| --- | --- | --- | --- | --- | --- |
| 1 | 2 | 1 (50) | 4 | 0.04 | 0.02 |
| 2 | 10 | 9 (90) | 22680 | 0.06 | 0.004 |
| 3 | 2 | 2 (100) | 12 | 0.11 | 0.01 |
| 4 | 2 | 1 (50) | 2 | 0.17 | -0.09 |
| 5 | 4 | 3 (75) | 64 | 0.25 | 0.07 |
| 6 | 19 | 2 (11) | 36 | 0.07 | -0.03 |
| 7 | 4 | 4 (100) | 480 | 0.36 | 0.03 |
| 8 | 5 | 3 (60) | 8 | 0.09 | -0.003 |
| 9 | 4 | 4 (100) | 18 | 1.37 | -0.59 |
| 10 | 5 | 1 (20) | 4 | 0.02 | -0.001 |
| 11 | 16 | 7 (44) | 192 | 0.13 | 0.03 |
| 12 | 7 | 6 (86) | 432 | 0.04 | -0.01 |
| 13 | 6 | 4 (67) | 64 | 0.03 | 0.01 |
| 14 | 6 | 3 (50) | 8 | 0.14 | 0.07 |
| 15 | 5 | 4 (80) | 1764 | 0.12 | 0.04 |
| 16 | 9 | 7 (78) | 1440 | 0.07 | -0.01 |
| 17 | 6 | 2 (33) | 4 | 0.13 | -0.05 |
| 18 | 7 | 7 (100) | 2880 | 0.10 | -0.01 |
| 19 | 8 | 8 (100) | 384 | 0.05 | 0.02 |
| 20 | 5 | 1 (20) | 2 | 0.05 | -0.02 |
| 21 | 9 | 8 (89) | 2916 | 0.17 | 0.03 |
| 22 | 19 | 11 (58) | 8192 | 0.02 | 0.01 |
| 23 | 11 | 6 (55) | 256 | 0.15 | -0.02 |

| SR ID | Number of study comparisons | Number (%) of study comparisons with multiplicity | Number of meta-analytic SMDs | Largest minus smallest meta-analytic SMD | Index minus median meta-analytic SMD<sup>a</sup> |
| --- | --- | --- | --- | --- | --- |
| 24 | 23 | 2 (9) | 27 | 0.16 | -0.01 |
| 25 | 23 | 19 (83) | 20155392<sup>b</sup> | 0.05 | 0.03 |
| 26 | 6 | 4 (67) | 16 | 0.58 | -0.29 |
| 27 | 20 | 14 (70) | 98304<sup>b</sup> | 0.03 | -0.01 |
| 28 | 5 | 4 (80) | 16 | 0.16 | -0.04 |
| 29 | 12 | 6 (50) | 288 | 0.02 | -0.001 |
| 30 | 16 | 1 (6) | 2 | 0.01 | 0.01 |
| 31 | 13 | 7 (54) | 384 | 0.20 | 0.03 |
| 32 | 10 | 6 (60) | 384 | 0.04 | -0.01 |
| 33 | 11 | 9 (82) | 3538944<sup>b</sup> | 0.15 | -0.02 |
| 34 | 16 | 10 (63) | 3456 | 0.03 | 0.01 |
| 35 | 13 | 7 (54) | 288 | 0.06 | -0.01 |
| 36 | 22 | 13 (59) | 110592<sup>b</sup> | 0.07 | -0.01 |
| 37 | 16 | 13 (81) | 16128000<sup>b</sup> | 0.09 | 0.01 |
| 38 | 4 | 4 (100) | 1152 | 0.11 | -0.01 |
| 39 | 5 | 1 (20) | 2 | 0.01 | -0.003 |
| Median (IQR) | 8 (5, 16) | 63% (50%, 83%) | 288 (12, 2880) | 0.09 (0.04, 0.15) | -0.003 (-0.01, 0.02) |

<sup>a</sup>Differences less than zero indicate that the index meta-analysis SMD is more favourable to the intervention compared with the median meta-analytic SMD.
<sup>b</sup>For these meta-analyses, we generated a sampling distribution of 5000 meta-analytic SMDs. Each meta-analytic SMD was created by randomly selecting (with equal probability) an effect estimate for inclusion from each study, and meta-analysing the chosen effects.

The median of the differences between the index meta-analytic SMD and the median of all its possible meta-analytic SMDs (i.e. the SMD expected when there is no selective inclusion) was −0.003 SD units (IQR −0.01 to 0.02; range −0.59 to 0.07). Meta-analysing these differences using a random-effects model yielded a pooled difference of 0.002 SD units (95% CI −0.01 to 0.01; I^2^=0%; Figure 3). Recalculating all possible meta-analytic SMDs using a fixed-effect model yielded nearly identical results (see Supplementary Figure S3; for results of all differences converted to ratios of odds ratios and sensitivity analyses, see Supplementary Figures S4-S9).

**Figure 3.**
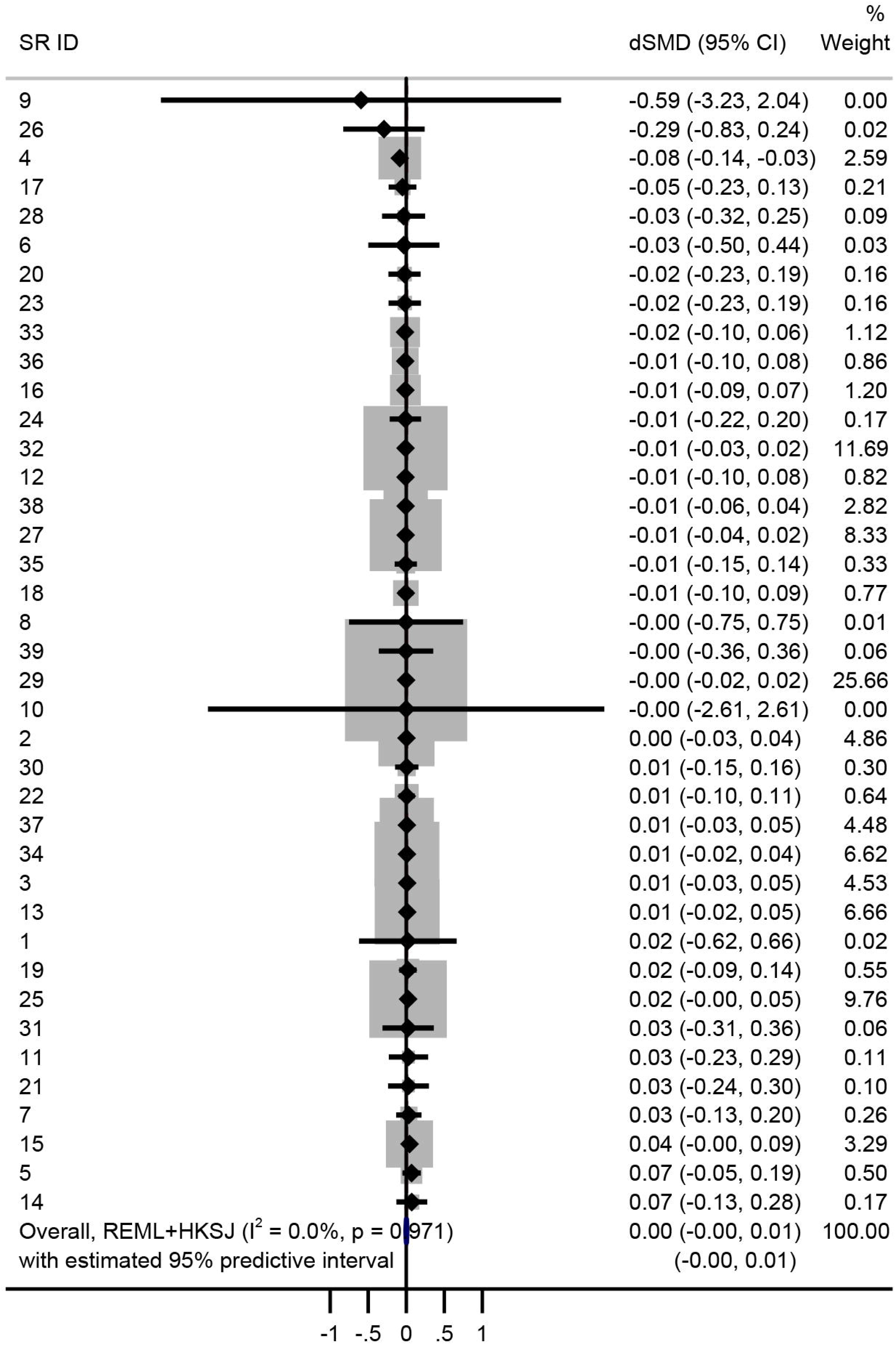
Meta-analysis of differences between the index meta-analytic SMD and median of all its possible meta-analytic SMDs (each calculated using the random-effects model). Differences less than zero indicate that the index meta-analysis SMD is more favourable to the intervention compared with the median meta-analytic SMD.

## 4. DISCUSSION

There was no evidence of selective inclusion of the most or least favourable reported study effect estimates by systematic reviewers in our sample of meta-analyses of food/diet-outcome associations. The PBI was not modified by outcome type for the index meta-analysis (continuous or non-continuous), funding source for the SR, or conflict of interests of systematic reviewers, but did differ depending on whether the review authors worked from a SR protocol/register entry. The PBI was robust to several assumptions in sensitivity analyses, including there being no evidence that the PBI estimates varied across the meta-analyses. On average, the index meta-analytic effect estimates were similar to what we would expect to see in meta-analyses generated when there was no selective inclusion.

Our investigation examined whether bias may be introduced through systematic reviewers’ decisions, which is in addition to bias that might arise from other sources (e.g. biased estimates in the constituent studies arising from methodological limitations; a biased subset of estimates reported by the study authors). While these other sources of bias could lead to biased meta-analysis estimates, which would be problematic in the context of a systematic review, they do not affect our investigation. The validity of our analyses relies upon identifying the set of study effect estimates that the systematic reviewers considered for inclusion in their meta-analyses; an issue we discuss further in Section 4.3.

### 4.1 Comparison with other studies

Our study results are largely similar to the only other study investigating bias due to selective inclusion of results^10^. In both studies, there was no clear evidence of selective inclusion of study estimates in the sample of meta-analyses evaluated, and importantly, in both studies the index meta-analytic effect estimates were similar, on average, to what we would expect to see in meta-analyses generated when there was no selective inclusion. The latter finding is explained by the fact that the range of possible meta-analytic SMDs calculated from the different combinations of study effect estimates was narrow; in the current study, the median difference (IQR) between the largest and smallest possible meta-analytic SMD was 0.09 SD units (IQR 0.04, 0.15), and in Page et al.^10^ it was similar (0.11 SD units; IQR 0.03, 0.19). Therefore, even in the event that reviewers did selectively include results, the potential for this to importantly impact the meta-analysis effect estimate was limited.

However, larger differences between the largest and smallest possible meta-analytic SMDs have been observed elsewhere. In a study evaluating multiplicity of effect estimates in study reports and its impact on meta-analysis results, Mayo-Wilson et al. observed a median difference between the largest and smallest possible meta-analytic SMD of 0.34 SD units^65^. In this circumstance, selective inclusion of results could impact meta-analysis effects; however, the authors did not explore selective inclusion of results in that study.

### 4.2 Explanations for study findings

There are several possible reasons why no evidence of selective inclusion of results was observed in this study. Despite infrequently pre-specifying or reporting methods for selecting results for inclusion in meta-analyses, the majority of systematic reviewers might have been following undisclosed decision rules that were based on some clinical or methodological rationale (i.e. not based on the statistical significance, magnitude or direction of effects). For example, systematic reviewers confronted with multiple effect estimates for the association between cheese intake and risk of prostate cancer, each adjusted for different sets of covariates, might have considered it reasonable to always select the most adjusted estimate available, without declaring in the Methods section that they followed this decision rule. Recommendations to report such selection methods only became available in sources such as the Cochrane Handbook^12^ and PRISMA 2020^66^ after all the SRs in this sample were published, which might explain the infrequent reporting of such methods.

Our observation that the PBI was more indicative of selective inclusion of results in SRs with a protocol/register entry than in SRs without a protocol/register entry was unanticipated. We assumed that review authors who publicly record their pre-specified methods may be more likely to specify methods for selecting results for inclusion in meta-analyses, and aware of the bias that can be introduced if this selection is made based on the nature of the result itself. However, of the 13 protocols/register entries, 11 were PROSPERO entries, and PROSPERO does not currently request authors pre-specify methods for selecting results for inclusion in meta-analyses. Therefore, simply registering a SR might not protect against selective inclusion at this stage. Furthermore, while the median number of eligible effect estimates was the same for studies included in SRs with a protocol/register entry and SRs without (median 2), the range of eligible effect estimates was narrower for the former (range 2-9 versus 2-21), which might have made it easier for SRs with a protocol/register entry to select the largest effect.

The similarity between the index and median meta-analytic effects may have been influenced by several factors. These include the percentage of studies with multiple effect estimates per meta-analysis, the extent to which the multiple effect estimates in a study varied in magnitude and direction, the weights that study effect estimates received in the meta-analysis or any combination of the above^10^. For example, if one adjusted estimate yields an OR of 1.87 and another yields an OR of 1.59, but the precision of the selected estimate was low and hence the study contributed little weight to the meta-analysis, selective inclusion of the most favourable OR is unlikely to affect the meta-analysis in an important way.

### 4.3 Strengths and weaknesses of the study

Our study has several strengths. We pre-specified our methods in a published protocol^11^, and have also specified any deviations from our protocol in Supplementary Table S1. We conducted a comprehensive search for SRs, and all records were screened by two authors independently to minimise errors in SR selection. Ranking of effect estimates for the PBI calculation was conducted by one author and verified by another.

There are also some limitations to our study. Data collection was conducted by two authors independently for only half of the SRs and their included studies. However, for the other half in which only one author collected data, we expect the number of data collection errors to be low because the collected data were verified for accuracy by another author. Our findings may be influenced by the set of study effect estimates that we considered eligible for inclusion in the meta-analyses, which might differ to what the systematic reviewers considered eligible. We used the methods reported in the SR protocols to select effect estimates; however, other selection methods may have been assumed but not documented at the protocol stage. We evaluated selective inclusion of results in the first reported meta-analysis in each systematic review. It is possible that the decision processes systematic reviewers use to select effects for inclusion differ across meta-analyses included within a review; however, this would seem unlikely. We only extracted data from the studies cited by the systematic reviewers. If other reports of studies (e.g. regulatory reports, conference abstracts) had been consulted but not cited by the systematic reviewers, our estimate of the true extent of multiplicity of results and the PBI might differ.

### 4.4 Future research

To date, only two studies (Page et al.^10^ and the current study) have investigated whether selective inclusion of study effect estimates occurs in meta-analyses in three areas (musculoskeletal conditions, mental health, nutrition), hence extending this type of investigation to other specialities will add to the evidence-base on the likelihood and impact of selective inclusion bias. Moreover, SRs that are funded by industry commonly report results favourable to the sponsor^67^, yet no study to date has examined the issue of selective inclusion in such SRs. Furthermore, it is worthwhile to explore bias due to selective inclusion of results in meta-analyses of harm outcomes, for which multiplicity of results is also a challenge^68, 69^. In addition, selective inclusion might be more likely to occur in reviews that have broad outcome domains, such as ‘patient health’, since within any study, there could be a large number of outcomes and effects that could be selected, with likely a wider range of effect estimates.

## 5. CONCLUSION

There was no evidence that systematic reviewers selectively included the most or least favourable reported study effect estimates in this sample of meta-analyses of the association between food/diet and health-related outcomes. Furthermore, the index meta-analytic effect estimates were similar, on average, to what we would expect to see in meta-analyses generated when there was no selective inclusion. Despite this, we encourage systematic reviewers to report whether they encountered multiplicity of results in the included studies, the methods they used to select effect estimates when multiple estimates were eligible for inclusion in a particular meta-analysis, and whether these selection methods were pre-specified. Doing so should help readers understand the risk of selective inclusion bias in the systematic review.

## DECLARATIONS

### Author contributions

All authors declare to meet the ICMJE conditions for authorship. MJP and JEM conceived the project. MJP, JEM, LB, ZD, SM, CMK and AF contributed to the design of the project. MJP, ZD, SM, and CMK screened articles for inclusion. RK, MJP, LB, ZD, SM, CMK, and EK collected data. RK and MJP analysed the data, adapting the analytic code written by JEM and AF for an earlier project. RK wrote the first draft of the manuscript, which was revised in conjunction with MJP and JEM. MJP and JEM drafted sections of the manuscript. All authors were involved in revising the article critically for important intellectual content. All authors approved the final version of the article. MJP is the guarantor of this work.

### Data availability statement

Data and analytic code are available on the Open Science Framework (https://osf.io/umk62/).

## HIGHLIGHTS

- Selective inclusion of results occurs when, for any particular primary study, multiple results are eligible for inclusion in a specific meta-analysis, and the result chosen for inclusion by the systematic reviewer is based on the nature of the result itself (e.g. the result’s P-value, magnitude or direction). Only one other study has investigated bias due to selective inclusion of results, in a sample of meta-analyses published between 2010 and 2012 and including randomised trials examining the effects of interventions for arthritis or depressive or anxiety disorders. It is unclear whether the findings of this prior study apply to other clinical conditions, outcome types (e.g. binary), study designs (e.g. non-randomised), and more recently published reviews.
- In a sample of 39 meta-analyses including 312 studies of nutrition research, there was no evidence that systematic reviewers selectively included the most, or least, favourable of the available study effect estimates. In addition, the index meta-analytic effect estimates were similar, on average, to what we would expect to see in a meta-analyses generated when there was no selective inclusion. The systematic reviews in our sample provided an opportunity to extend the previous investigation of bias due to selective inclusion of results, because these reviews often include study designs beyond randomised trials, different outcome types, and include results from multiple analyses that attempt to adjust for potential confounding variables (in the case of non-randomised studies).
- We recommend that systematic reviewers report the methods used to select effect estimates to include in meta-analyses, which can help readers understand the risk of selective inclusion bias in the systematic reviews.

## Data Availability

Data and analytic code are available on the Open Science Framework (https://osf.io/umk62/).

## REFERENCES

1. Kurien VTV, Shamsuddeen S, Mahitha M, Rasheed DS. Evidence-based decision-making. Journal of Head & Neck Physicians and Surgeons 2022;10(1):48.

2. Schünemann HJ, Wiercioch W, Etxeandia I, et al. Guidelines 2.0: systematic development of a comprehensive checklist for a successful guideline enterprise. CMAJ 2014;186(3):E123–42.

3. Mayo-Wilson E, Fusco N, Li T, Hong H, Canner JK, Dickersin K. Multiple outcomes and analyses in clinical trials create challenges for interpretation and research synthesis. J Clin Epidemiol 2017;86:39–50.

4. Page MJ, McKenzie JE, Forbes A. Many scenarios exist for selective inclusion and reporting of results in randomized trials and systematic reviews. J Clin Epidemiol 2013;66(5):524–37.

5. Kanukula R, McKenzie JE, Bero L, et al. Methods used to select results to include in meta-analyses of nutrition research: A meta-research study. J Clin Epidemiol 2022;142:171–183.

6. López-López JA, Page MJ, Lipsey MW, Higgins JPT. Dealing with effect size multiplicity in systematic reviews and meta-analyses. Res Synth Methods 2018;9(3):336–351.

7. Konstantopoulos S. Fixed effects and variance components estimation in three-level meta-analysis. Res Synth Methods 2011;2(1):61–76.

8. Hedges LV, Tipton E, Johnson MC. Robust variance estimation in meta-regression with dependent effect size estimates. Res Synth Methods 2010;1(1):39–65.

9. Page MJ, McKenzie JE, Chau M, Green SE, Forbes A. Methods to select results to include in meta-analyses deserve more consideration in systematic reviews. J Clin Epidemiol 2015;68(11):1282–1291.

10. Page MJ, Forbes A, Chau M, Green SE, McKenzie JE. Investigation of bias in meta-analyses due to selective inclusion of trial effect estimates: empirical study. BMJ Open 2016;6(4):e011863.

11. Page MJ, Bero L, Kroeger CM, et al. Investigation of risk of bias due to unreported and selectively included results in meta-analyses of nutrition research: the ROBUST study protocol. F1000Research 2019;8:1760.

12. Higgins JP, Thomas J, Chandler J, et al. Cochrane Handbook for Systematic Reviews of Interventions. John Wiley & Sons; 2019.

13. Harris PA, Taylor R, Minor BL, et al. The REDCap consortium: Building an international community of software platform partners. Journal of Biomedical Informatics 2019;95:103208.

14. Page MJ, McKenzie JE, Green SE, Forbes AB. An empirical investigation of the potential impact of selective inclusion of results in systematic reviews of interventions: study protocol. Syst Rev 2013;2:21.

15. Efron B, Tibshirani R. An Introduction to the Bootstrap. New York, NY: Chapman & HallHall. 1993;

16. Hartung J, Knapp G. A refined method for the metaLanalysis of controlled clinical trials with binary outcome. Statistics in Medicine 2001;20(24):3875–3889.

17. Sidik K, Jonkman JN. A simple confidence interval for metaLanalysis. Statistics in Medicine 2002;21(21):3153–3159.

18. Higgins JP, Thompson SG, Deeks JJ, Altman DG. Measuring inconsistency in meta-analyses. BMJ 2003;327(7414):557–560.

19. Deeks JJ, Higgins JP, Altman D. Chapter 10: Analysing data and undertaking meta-analyses. In: Higgins JP, Thomas J, Chandler J, et al, eds. Cochrane Handbook for Systematic Reviews of Interventions. John Wiley & Sons; 2019.

20. Herbison P, Robertson MC, McKenzie JE. Do alternative methods for analysing count data produce similar estimates? Implications for meta-analyses. Syst Rev 2015;4(1):163.

21. Chinn S. A simple method for converting an odds ratio to effect size for use in metaLanalysis. Statistics in Medicine 2000;19(22):3127–3131.

22. Stata Statistical Software: Release 16. College Station, TX: StataCorp LLC. Version 16. StataCorp.; 2019.

23. Ayoub-Charette S, Liu Q, Khan TA, et al. Important food sources of fructose-containing sugars and incident gout: a systematic review and meta-analysis of prospective cohort studies. BMJ Open 2019;9(5):e024171.

24. Cheng S, Zheng Q, Ding G, Li G. Mediterranean dietary pattern and the risk of prostate cancer: A meta-analysis. Medicine 2019;98(27):e16341.

25. Choo VL, Viguiliouk E, Mejia SB, et al. Food sources of fructose-containing sugars and glycaemic control: systematic review and meta-analysis of controlled intervention studies. BMJ 2018;363:k4644.

26. de Magalhães Cunha C, Costa PR, de Oliveira LP, Valterlinda AdO, Pitangueira JC, Oliveira AM. Dietary patterns and cardiometabolic risk factors among adolescents: systematic review and meta-analysis. British Journal of Nutrition 2018;119(8):859–879.

27. Eaton JC, RothpletzLPuglia P, Dreker MR, et al. Effectiveness of provision of animalLsource foods for supporting optimal growth and development in children 6 to 59 months of age. Cochrane Database of Systematic Reviews 2019;2(2):CD012818.

28. George ES, Marshall S, Mayr HL, et al. The effect of high-polyphenol extra virgin olive oil on cardiovascular risk factors: A systematic review and meta-analysis. Critical Reviews in Food Science and Nutrition 2019;59(17):2772–2795.

29. Ghaedi E, Mohammadi M, Mohammadi H, et al. Effects of a Paleolithic diet on cardiovascular disease risk factors: a systematic review and meta-analysis of randomized controlled trials. Advances in Nutrition 2019;10(4):634–646.

30. Haider LM, Schwingshackl L, Hoffmann G, Ekmekcioglu C. The effect of vegetarian diets on iron status in adults: A systematic review and meta-analysis. Critical Reviews in Food Science and Nutrition 2018;58(8):1359–1374.

31. Hou R, Wei J, Hu Y, et al. Healthy dietary patterns and risk and survival of breast cancer: a meta-analysis of cohort studies. Cancer Causes & Control 2019;30(8):835–846.

32. Huang Y, Zheng S, Wang T, Yang X, Luo Q, Li H. Effect of oral nut supplementation on endothelium-dependent vasodilation–a meta-analysis. Vasa 2018;47(3):203–207.

33. Iguacel I, Miguel-Berges ML, Gómez-Bruton A, Moreno LA, Julián C. Veganism, vegetarianism, bone mineral density, and fracture risk: a systematic review and meta-analysis. Nutrition Review. 2019;77(1):1–18.

34. Kang K, Sotunde OF, Weiler HA. Effects of milk and milk-product consumption on growth among children and adolescents aged 6–18 years: a meta-analysis of randomized controlled trials. Advances in Nutrition 2019;10(2):250–261.

35. Kibret KT, Chojenta C, Gresham E, Tegegne TK, Loxton D. Maternal dietary patterns and risk of adverse pregnancy (hypertensive disorders of pregnancy and gestational diabetes mellitus) and birth (preterm birth and low birth weight) outcomes: a systematic review and meta-analysis. Public Health Nutrition 2019;22(3):506–520.

36. Kodama S, Horikawa C, Fujihara K, et al. Relationship between intake of fruit separately from vegetables and triglycerides-A meta-analysis. Clinical Nutrition ESPEN 2018;27:53–58.

37. Kojima G, Avgerinou C, Iliffe S, Walters K. Adherence to Mediterranean diet reduces incident frailty risk: systematic review and metaLanalysis. Journal of the American Geriatrics Society 2018;66(4):783–788.

38. Larsson SC, Drca N, Jensen-Urstad M, Wolk A. Chocolate consumption and risk of atrial fibrillation: two cohort studies and a meta-analysis. American Heart Journal 2018;195:86–90.

39. Li R, Yu K, Li C. Dietary factors and risk of gout and hyperuricemia: a meta-analysis and systematic review. Asia Pacific Journal of Clinical Nutrition 2018;27(6):1344–1356.

40. Li W, Ruan W, Peng Y, Wang D. Soy and the risk of type 2 diabetes mellitus: A systematic review and meta-analysis of observational studies. Diabetes Research and Clinical Practice 2018;137:190–199.

41. Li L, Lietz G, Seal C. Buckwheat and CVD risk markers: A systematic review and meta-analysis. Nutrients 2018;10(5):619.

42. Lopez PD, Cativo EH, Atlas SA, Rosendorff C. The effect of vegan diets on blood pressure in adults: a meta-analysis of randomized controlled trials. The American Journal of Medicine 2019;132(7):875–883.

43. Maki KC, Palacios OM, Koecher K, et al. The relationship between whole grain intake and body weight: results of meta-analyses of observational studies and randomized controlled trials. Nutrients 2019;11(6):1245.

44. Matía-Martín P, Torrego-Ellacuría M, Larrad-Sainz A, Fernández-Pérez C, Cuesta-Triana F, Rubio-Herrera MÁ. Effects of milk and dairy products on the prevention of osteoporosis and osteoporotic fractures in Europeans and non-Hispanic Whites from North America: a systematic review and updated meta-analysis. Advances in Nutrition 2019;10(suppl_2):S120–S143.

45. Mena-Sánchez G, Becerra-Tomás N, Babio N, Salas-Salvadó J. Dairy product consumption in the prevention of metabolic syndrome: a systematic review and meta-analysis of prospective cohort studies. Advances in Nutrition 2019;10(suppl_2):S144–S153.

46. Milajerdi A, Namazi N, Larijani B, Azadbakht L. The association of dietary quality indices and cancer mortality: a systematic review and meta-analysis of cohort studies. Nutrition and Cancer 2018;70(7):1091–1105.

47. Mishali M, Prizant-Passal S, Avrech T, Shoenfeld Y. Association between dairy intake and the risk of contracting type 2 diabetes and cardiovascular diseases: a systematic review and meta-analysis with subgroup analysis of men versus women. Nutrition Reviews 2019;77(6):417–429.

48. Mohseni R, Abbasi S, Mohseni F, Rahimi F, Alizadeh S. Association between dietary inflammatory index and the risk of prostate cancer: a meta-analysis. Nutrition and Cancer 2019;71(3):359–366.

49. Namazi N, Larijani B, Azadbakht L. Dietary inflammatory index and its association with the risk of cardiovascular diseases, metabolic syndrome, and mortality: a systematic review and meta-analysis. Hormone and Metabolic Research 2018;50(05):345–358.

50. Picasso MC, Lo-Tayraco JA, Ramos-Villanueva JM, Pasupuleti V, Hernandez AV. Effect of vegetarian diets on the presentation of metabolic syndrome or its components: A systematic review and meta-analysis. Clinical Nutrition 2019;38(3):1117–1132.

51. Qin Z-Z, Xu J-Y, Chen G-C, Ma Y-X, Qin L-Q. Effects of fatty and lean fish intake on stroke risk: a meta-analysis of prospective cohort studies. Lipids in Health and Disease 2018;17(1):1–7.

52. Rees K, Takeda A, Martin N, et al. Mediterranean style diet for the primary and secondary prevention of cardiovascular disease. Cochrane Database of Systematic Reviews 2019;3(3):CD009825.

53. Ren Y, Liu Y, Sun X-Z, et al. Chocolate consumption and risk of cardiovascular diseases: a meta-analysis of prospective studies. Heart 2019;105(1):49–55.

54. Shab-Bidar S, Golzarand M, Hajimohammadi M, Mansouri S. A posteriori dietary patterns and metabolic syndrome in adults: a systematic review and meta-analysis of observational studies. Public Health Nutrition 2018;21(9):1681–1692.

55. Shafiei F, Salari-Moghaddam A, Larijani B, Esmaillzadeh A. Adherence to the Mediterranean diet and risk of depression: a systematic review and updated meta-analysis of observational studies. Nutrition Reviews 2019;77(4):230–239.

56. Teoh SL, Lai NM, Vanichkulpitak P, Vuksan V, Ho H, Chaiyakunapruk N. Clinical evidence on dietary supplementation with chia seed (Salvia hispanica L.): a systematic review and meta-analysis. Nutrition Reviews 2018;76(4):219–242.

57. Voon PT, Lee ST, Ng TKW, et al. Intake of palm olein and lipid status in healthy adults: A meta-analysis. Advances in Nutrition 2019;10(4):647–659.

58. Wang L, Liu C, Zhou C, et al. Meta-analysis of the association between the dietary inflammatory index (DII) and breast cancer risk. European Journal of Clinical Nutrition 2019;73(4):509–517.

59. Xiao Y, Ke Y, Wu S, et al. Association between whole grain intake and breast cancer risk: a systematic review and meta-analysis of observational studies. Nutrition Journal 2018;17(1):1–12.

60. Xu Y, Yang J, Du L, Li K, Zhou Y. Association of whole grain, refined grain, and cereal consumption with gastric cancer risk: A metaLanalysis of observational studies. Food Science & Nutrition 2019;7(1):256–265.

61. Zhang Z, Chen G-C, Qin Z-Z, Tong X, Li D-P, Qin L-Q. Poultry and fish consumption in relation to total cancer mortality: a meta-analysis of prospective studies. Nutrition and Cancer 2018;70(2):204–212.

62. Bermejo LM, López-Plaza B, Santurino C, Cavero-Redondo I, Gómez-Candela C. Milk and dairy product consumption and bladder cancer risk: a systematic review and meta-analysis of observational studies. Advances in Nutrition 2019;10(suppl_2):S224–S238.

63. Malmir H, Saneei P, Larijani B, Esmaillzadeh A. Adherence to Mediterranean diet in relation to bone mineral density and risk of fracture: a systematic review and meta-analysis of observational studies. European Journal of Nutrition 2018;57(6):2147–2160.

64. Musa-Veloso K, Poon T, Harkness LS, O’Shea M, Chu Y. The effects of whole-grain compared with refined wheat, rice, and rye on the postprandial blood glucose response: a systematic review and meta-analysis of randomized controlled trials. The American Journal of Clinical Nutrition 2018;108(4):759–774.

65. Mayo-Wilson E, Li T, Fusco N, et al. Cherry-picking by trialists and meta-analysts can drive conclusions about intervention efficacy. J Clin Epidemiol 2017;91:95–110.

66. Page MJ, McKenzie JE, Bossuyt PM, et al. The PRISMA 2020 statement: an updated guideline for reporting systematic reviews. BMJ 2021;372:n71.

67. Ebrahim S, Bance S, Athale A, Malachowski C, Ioannidis JP. Meta-analyses with industry involvement are massively published and report no caveats for antidepressants. J Clin Epidemiol 2016;70:155–163.

68. Mayo-Wilson E, Fusco N, Li T, et al. Harms are assessed inconsistently and reported inadequately Part 1: Systematic adverse events. J Clin Epidemiol 2019;113:20–27.

69. Mayo-Wilson E, Fusco N, Li T, et al. Harms are assessed inconsistently and reported inadequately Part 2: Nonsystematic adverse events. J Clin Epidemiol 2019;113:11–19.

