## Supplementary for "Investigation of bias due to selective inclusion of study effect estimates in meta-analyses of nutrition research"

**Supplementary files**

**Supplementary Table S1. Modifications from the ROBUST study protocol**

| **Original plan** | **Revised plan** | **Reason for modification** |
| --- | --- | --- |
| We will seek a sample of SRs with meta-analysis meeting the following criteria:  • includes studies of people of any ages (i.e. infants, children, adolescents, adults or elderly people) and backgrounds in the generally healthy population, including pregnant and breastfeeding women and people with common diet-related risk factors such as being overweight or having high blood pressure | We will seek a sample of SRs with meta-analysis meeting the following criteria:  • includes studies that enrolled (a) people of any age or background who were generally healthy, (b) a mixture of generally healthy people and people with diet-related risk factors (e.g. overweight, high blood pressure) or a particular health condition (e.g. type II diabetes or cardiovascular disease), or (c) people with non-specified health status; | During screening of full text articles, we discovered that many otherwise eligible SRs did not state the health status of participants in included studies, or included a mix of participants who were healthy or those who had a particular clinical condition. After screening 1,500 abstracts and retrieving 149 full text articles, 44 SRs were considered to meet the eligibility criteria; however, on closer inspection, only 7 SRs included studies restricted to “healthy people”, 12 SRs included a mixture of studies of healthy people and people with a health condition (e.g. type II diabetes), and in 25 SRs the health status of participants was not clear (e.g. the authors said they included studies of “adults” and the characteristics of included studies presented provided no information on health status or risk factors). We have no reason to believe that SRs meeting the original inclusion criteria would be systematically different from SRs meeting the revised criteria. Therefore, rather than continue screening abstracts meeting the more stringent, original criteria, we decided to include SRs meeting the less stringent, revised criteria. |
| We will include meta-analyses of binary outcomes and meta-analyses of continuous outcomes. | We will include meta-analyses of non-continuous outcomes (e.g. binary, count, time-to-event) and meta-analyses of continuous outcomes. | During screening of articles we decided to expand eligibility from meta-analyses of “binary” to meta-analyses of “non-continuous” outcomes given the types of multiplicity affecting these meta-analyses are similar. |
| No plan for dealing with the scenario whereby screening of full text articles results in more than 25 SRs of one type (with binary or continuous outcomes). | If, during the screening process the total number of eligible SRs identified exceeded the target of 25 SRs with a meta-analysis of a non-continuous outcome and 25 SRs with a meta-analysis of a continuous outcome, we will randomly sample 25 SRs of each type. | By the time we had identified 25 eligible systematic reviews with meta-analysis of a continuous outcome, we had identified 74 eligible SRs with meta-analysis of a non-continuous outcome. Rather than include all SRs with meta-analysis of a non-continuous outcome in the study, we decided to randomly select 25 SRs. |
| Following piloting, two investigators will independently collect data from all remaining index meta-analyses and included studies. | Following piloting, two investigators will independently collect data on half of the remaining index meta-analyses and their included studies, while one investigator will collect data on the other half of the remaining index meta-analyses and their included studies. We will randomly select the SRs which will have their data collected by two investigators. | We decided to reduce the workload for investigators responsible for collecting data. |
| We will select a random sample of 50 SRs. From each SR, one investigator will select one meta-analysis of a binary or continuous outcome for assessment, stratifying the selection so that the final sample includes 25 binary and 25 continuous outcome meta-analyses. We will extract data from all SRs selected, regardless of the number of included studies in the index meta-analysis. | We will select a random sample of 50 SRs, 25 of which will have a non-continuous index meta-analysis and 25 of which will have a continuous index meta-analysis. However, we will only extract data from SRs with less than 20 studies included in the index meta-analysis. | We decided to reduce the workload for investigators responsible for collecting data. |
| No plan for handling subgroup effect estimates in studies. | We will only extract subgroup effect estimates from studies if (a) they were the most compatible with the index meta-analysis (e.g. if the index meta-analysis is defined as “cancer mortality in women”, we will only extract subgroup estimates of cancer mortality for women); (b) they were the only effect estimates available in a paper and both were compatible with the index meta-analysis (e.g. if the index meta-analysis is defined as “cancer mortality” and study investigators report an estimate for men and an estimate for women, but not the combined estimate, we will extract both subgroup estimates); or (c) the systematic reviewers included subgroup effect estimates in the index meta-analysis. We will not extract subgroup effect estimates from studies if none of the 3 criteria above are met, and if an estimate based on the total sample of participants was available in the paper. | Subgroup effect estimates are less precise and less generalisable than effect estimates based on the total sample. For this reason, selecting an effect estimate based on the total sample (when available) ahead of the subgroup effect estimates should not be considered selective inclusion of results by reviewers. |
| In our primary analysis of meta-analyses of continuous outcomes, we will convert all study effect estimates to SMDs to allow us to calculate the PBI in circumstances where multiple effect estimates were available for the same outcome domain, but measured on different scales. However, there is not necessarily a one-to-one relationship between the rank positions of effect estimates based on the mean difference and SMD (because the SMD additionally depends on the pooled standard deviation). Therefore, in a sensitivity analysis we will calculate the PBI based on the rank positions of the mean difference for the subset of study effect estimates that were measured on the same scale as the effect estimate included in the index meta-analysis. This will allow us to assess more accurately whether systematic reviewers had selectively included study effect estimates based on the magnitude of the mean difference in raw measurement scale units. | We will not undertake this sensitivity analysis. | The proposed sensitivity analysis was unnecessary. All meta-analyses of continuous outcomes (including those that used the SMD as the effect measure) analysed outcomes for which all study effect estimates were in the same measurement units. Therefore, we were able to rank mean differences rather than SMDs. |

**Supplementary Table S2: Search Strategies**

| **Database** | **Search strategy** |
| --- | --- |
| PubMed | 1. Nutritional Sciences[mh] OR Nutritional Physiological Phenomena[mh] OR Nutrition Assessment[mh] OR Nutrition Therapy[mh] OR Nutrition Policy[mh] OR Nutritional and Metabolic Diseases[mh] OR nutrition*[tiab] OR diet[tiab] OR feeding[tiab] OR dietary[tiab] OR breastfeed*[tiab] OR breast feed*[tiab] OR lactation[tiab] OR bottle feed*[tiab] OR complementary feeding[tiab] OR weaning[tiab] OR enteral[tiab] OR parenteral[tiab] OR Feeding Methods[mh] OR nutritional status[tiab] OR overweight[tiab] OR obese[tiab] OR obesity[tiab] OR overnutrition[tiab] OR over nutrition[tiab] OR undernourished[tiab] OR overnourished[tiab] OR wasted[tiab] OR wasting[tiab] OR stunting[tiab] OR stunted[tiab] OR underweight[tiab] OR undernutrition[tiab] OR under nutrition[tiab] OR body weight[tiab] OR anthropometry[tiab] OR Body Weights and Measures[mh] OR growth monitoring[tiab] OR food[tiab] OR food labelling[mh] OR food assistance[mh] OR supplementary feeding[tiab] OR diet therapy[mh] OR food and beverages[mh] OR vegetable*[tiab] OR fruit*[tiab] OR meat[tiab] OR dairy[tiab] OR dietary fat*[tiab] OR starch*[tiab] OR cereal[tiab] OR food-drug interactions[mh] OR food supply[mh] OR feeding behavio*[tiab] OR eating behavio*[tiab] OR food pattern*[tiab] OR food hypersensitivity[mh] OR food deprivation[mh] OR food, organic [mh] OR micronutrient*[tiab] OR vitamin*[tiab] OR thiamin[tiab] OR riboflavin[tiab] OR niacin[tiab] OR pantothenic acid[tiab] OR pyridoxine[tiab] OR pyridoxal[tiab] OR pyridoxamine[tiab] OR biotin[tiab] OR folic acid[tiab] OR folate[tiab]OR cyanocobalamin[tiab] OR choline[tiab] OR retinol[tiab] OR ascorbic acid[tiab] OR tocopherol[tiab] OR carotenoids[tiab] OR carotene[tiab] OR cryptoxanthin[tiab] OR lutein[tiab] OR lycopene[tiab] OR zeaxanthin[tiab] OR minerals[tiab] OR calcium[tiab] OR chloride[tiab] OR magnesium[tiab] OR phosphorus[tiab] OR potassium[tiab] OR sodium[tiab] OR iron[tiab] OR sulphur[tiab] OR trace element*[tiab] OR boron[tiab] OR cobalt[tiab] OR chromium[tiab] OR copper[tiab] OR fluoride[tiab] OR iodine[tiab] OR iron[tiab] OR manganese[tiab] OR molybdenum[tiab] OR selenium[tiab] OR zinc[tiab] OR trace metal*[tiab] OR macronutrient*[tiab] OR carbohydrate*[tiab] OR dietary protein*[tiab] OR saturated fat*[tiab] OR unsaturated fat*[tiab] OR mono unsaturated fat*[tiab] OR monounsaturated fat*[tiab] OR poly unsaturated fat*[tiab] OR polyunsaturated fat*[tiab] OR trans fat*[tiab] OR dietary fibre[tiab] OR dietary fiber[tiab] OR dietary salt[tiab] OR table salt[tiab] OR soft drink[tiab] OR fruit juice[tiab] OR vegetable juice[tiab] OR milk[tiab] OR tea[tiab] OR coffee[tiab] OR energy drink*[tiab] OR carbonated beverage*[tiab] OR carbonated drink*[tiab] OR prebiotics[tiab] OR probiotics[tiab] OR glycemic load[tiab] OR glycemic index[tiab] OR glycaemic load[tiab] OR glycaemic index[tiab] OR calories[tiab] OR kilocalories[tiab] OR kilojoules[tiab] OR caloric intake[tiab] OR energy intake[tiab] 2. meta-analysis[pt] OR meta-analy*[ti] 3. ("2018/01/01"[PDAT] : "2019/06/30"[PDAT]) 4. 1 AND 2 AND 3 |
| Epistemonikos | 1. (advanced_title_en:(nutri* OR diet*) OR advanced_abstract_en:(nutri* OR diet*)) [Filters: protocol=no, min_year=2018, max_year=2019] |

| **Supplementary Table S3: Data collection form** |
| --- |

|  | **#** | **Variable / Field Name** | **Field Label**  ***Field Note*** | **Field Attributes (Field Type, Validation, Choices, Calculations, etc.)** |
| --- | --- | --- | --- | --- |
| Instrument: **SR protocol characteristics** (sr_protocol_characteristics) | | | | |
|  | 1 | record_id | Record ID: | text |
|  | 2 | sr_id | SR ID (stated on page 1 of PDF): | text |
|  | 3 | initials | Enter initials of data collector: | text |
|  | 4 | index_ma | Specify the index meta-analysis outcome (stated on pg 1 of the PDF): | text |
|  | 5 | protocol_avail | Is a protocol or registration record available for the SR? | radio 1 \| Both a protocol and registration record are available; 2 \| Only a protocol is available; 3 \| Only a registration record is available; 4 \| Neither are available |
|  | 6 | srp_year  Show the field ONLY if:  [protocol_avail] = '1' or [protocol_avail] = '2' | Specify the year of publication of the protocol: | text |
|  | 7 | regist_year  Show the field ONLY if:  [protocol_avail] = '1' or [protocol_avail] = '3' | Specify the year the SR was first registered: | text |
|  | 8 | srp_e  Show the field ONLY if:  [protocol_avail] = '1' or [protocol_avail] = '2' or [protocol_avail] = '3' | Section Header: *Eligibility criteria and decision rules*  Review authors pre-specified...(select ALL of the following that apply): | checkbox 1 \| srp_e___1 \| ...eligible measurement instruments for [index_ma]; 2 \| srp_e___2 \| ...eligible definitions/diagnostic criteria for [index_ma]; 3 \| srp_e___3 \| ...eligible cut-points on an instrument for [index_ma]; 4 \| srp_e___4 \| ...eligible time points for [index_ma]; 5 \| srp_e___5 \| ...eligible interventions/exposures; 6 \| srp_e___6 \| ...whether final values or change from baseline values (or both) for [index_ma] were eligible; 7 \| srp_e___7 \| ...which analysis samples for [index_ma] were eligible (e.g. intention-to-treat, per-protocol or as-treated); 8 \| srp_e___8 \| ...whether unadjusted or covariate-adjusted analyses (or both) for [index_ma] were eligible; 9 \| srp_e___9 \| ...whether period or paired analyses (or both) for [index_ma] were eligible (from crossover trials); 10 \| srp_e___10 \| ...which information sources were eligible (e.g. journal article, trials results register) |
|  | 9 | srp_dr  Show the field ONLY if:  [protocol_avail] = '1' or [protocol_avail] = '2' or [protocol_avail] = '3' | Review authors pre-specified...(select ALL of the following that apply): | checkbox 1 \| srp_dr___1 \| ...a decision rule to handle results arising from multiple measurement instruments for [index_ma]; 2 \| srp_dr___2 \| ...a decision rule to handle results arising from multiple definitions/diagnostic criteria for [index_ma]; 3 \| srp_dr___3 \| ...a decision rule to handle results arising from multiple cut-points on an instrument for [index_ma]; 4 \| srp_dr___4 \| ...a decision rule to handle results arising from multiple time points for [index_ma]; 5 \| srp_dr___5 \| ...a decision rule to handle results arising from multiple interventions/exposures; 6 \| srp_dr___6 \| ...a decision rule to handle results arising from both final values and change from baseline values for [index_ma]; 7 \| srp_dr___7 \| ...a decision rule to handle results arising from multiple analysis samples (e.g. intention-to-treat, per-protocol or as-treated) for [index_ma]; 8 \| srp_dr___8 \| ...a decision rule to handle results arising from multiple unadjusted or covariate-adjusted analyses for [index_ma]; 9 \| srp_dr___9 \| ...a decision rule to handle results arising from period and paired analyses for [index_ma] in crossover trials; 10 \| srp_dr___10 \| ...a decision rule to handle results arising from multiple information sources (e.g. journal article, trials results register); 11 \| srp_dr___11 \| ...a decision rule to handle results arising from overlapping samples of participants (e.g. subgroup of men of all ages and subgroup of older adults which includes men); 12 \| srp_dr___12 \| ...a decision rule to handle results arising from another source of multiplicity |
|  | 10 | srp_e_scale_txt  Show the field ONLY if:  [srp_e(1)] = '1' | Specify the eligible measurement instruments for [index_ma]: | notes |
|  | 11 | srp_dr_scale_txt  Show the field ONLY if:  [srp_dr(1)] = '1' | Specify the decision rule to handle results arising from multiple measurement instruments for [index_ma]: | notes |
|  | 12 | srp_e_event_txt  Show the field ONLY if:  [srp_e(2)] = '1' | Specify the eligible definitions/diagnostic criteria for [index_ma]: | notes |
|  | 13 | srp_dr_event_txt  Show the field ONLY if:  [srp_dr(2)] = '1' | Specify the decision rule to handle results arising from multiple definitions/diagnostic criteria for [index_ma]: | notes |
|  | 14 | srp_e_cutpoint_txt  Show the field ONLY if:  [srp_e(3)] = '1' | Specify the eligible cut-points on an instrument for [index_ma]: | notes |
|  | 15 | srp_dr_cutpoint_txt  Show the field ONLY if:  [srp_dr(3)] = '1' | Specify the decision rule to handle results arising from multiple cut-points on an instrument for [index_ma]: | notes |
|  | 16 | srp_e_time_txt  Show the field ONLY if:  [srp_e(4)] = '1' | Specify the eligible time points for [index_ma]: | notes |
|  | 17 | srp_dr_time_txt  Show the field ONLY if:  [srp_dr(4)] = '1' | Specify the decision rule to handle results arising from multiple time points for [index_ma]: | notes |
|  | 18 | srp_e_group_txt  Show the field ONLY if:  [srp_e(5)] = '1' | Specify the eligible interventions/exposures (incl. comparators): | notes |
|  | 19 | srp_dr_group_txt  Show the field ONLY if:  [srp_dr(5)] = '1' | Specify the decision rule to handle results arising from multiple interventions/exposures: | notes |
|  | 20 | srp_e_fvcs_txt  Show the field ONLY if:  [srp_e(6)] = '1' | Specify which values (i.e. final, change from baseline or both) for [index_ma] were eligible: | notes |
|  | 21 | srp_dr_fvcs_txt  Show the field ONLY if:  [srp_dr(6)] = '1' | Specify the decision rule to handle results arising from both final and change from baseline values for [index_ma]: | notes |
|  | 22 | srp_e_itt_txt  Show the field ONLY if:  [srp_e(7)] = '1' | Specify which analysis samples (e.g. intention-to-treat, per-protocol) for [index_ma] were eligible: | notes |
|  | 23 | srp_dr_itt_txt  Show the field ONLY if:  [srp_dr(7)] = '1' | Specify the decision rule to handle results arising from multiple analysis samples (e.g. intention-to-treat, per-protocol) for [index_ma]: | notes |
|  | 24 | srp_e_unadj_txt  Show the field ONLY if:  [srp_e(8)] = '1' | Specify which unadjusted and covariate-adjusted analyses for [index_ma] were eligible: | notes |
|  | 25 | srp_dr_unadj_txt  Show the field ONLY if:  [srp_dr(8)] = '1' | Specify the decision rule to handle results arising from multiple unadjusted and covariate-adjusted analyses for [index_ma]: | notes |
|  | 26 | srp_e_cross_txt  Show the field ONLY if:  [srp_e(9)] = '1' | Specify which analyses in crossover trials (e.g. first period, second period, paired) were eligible: | notes |
|  | 27 | srp_dr_cross_txt  Show the field ONLY if:  [srp_dr(9)] = '1' | Specify the decision rule to handle results arising from period and paired analyses in crossover trials: | notes |
|  | 28 | srp_e_source_txt  Show the field ONLY if:  [srp_e(10)] = '1' | Specify which information sources (e.g. journal article, trials results register) were eligible: | notes |
|  | 29 | srp_dr_source_txt  Show the field ONLY if:  [srp_dr(10)] = '1' | Specify the decision rule to handle results arising from multiple information sources (e.g. journal article, trials results register): | notes |
|  | 30 | srp_dr_overlap_txt  Show the field ONLY if:  [srp_dr(11)] = '1' | Specify the decision rule to handle results arising from overlapping samples of participants: | Notes |
|  | 31 | srp_dr_other_txt  Show the field ONLY if:  [srp_dr(12)] = '1' | Specify the decision rule(s) to handle results arising from other sources of multiplicity for [index_ma]: | Notes |
|  | 32 | srp_notes | Notes | Notes |
|  | 33 | sr_protocol_characteristics_complete | Section Header: *Form Status*  Complete? | Dropdown 0 \| Incomplete; 1 \| Unverified; 2 \| Complete |
| Instrument: **SR characteristics** (sr_characteristics) | | | | |
|  | 34 | sr_title | Specify the title of the systematic review: | Text |
|  | 35 | sr_journal | Specify the journal: | Text |
|  | 36 | sr_year | Specify the year of publication: | Text |
|  | 37 | sr_country | Specify the country of the corresponding author of the SR: | Text |
|  | 38 | sr_funding | What was the source of funding for the SR? | Radio 1 \| Non-profit; 2 \| For-profit; 3 \| Mixed; 4 \| No funding; 5 \| Not reported |
|  | 39 | for_profit_org  Show the field ONLY if:  [sr_funding] = '2' or [sr_funding] = '3' | What type of for-profit organisation funded the SR? | Radio 1 \| Food industry; 2 \| Other industry |
|  | 40 | for_profit_org_txt  Show the field ONLY if:  [sr_funding] = '2' or [sr_funding] = '3' | Record the name of the for-profit funder: | Text |
|  | 41 | sr_coi | Did any of the systematic reviewers disclose financial conflicts of interest? | Radio 1 \| Conflict of interest present (i.e. at least one systematic reviewer reported a financial conflict of interest of any type, excluding current study funding or industry employment); 2 \| No conflict of interest (i.e. all systematic reviewers stated they had no conflicts); 3 \| Missing (i.e. no disclosure statement) |
|  | 42 | sr_coi_txt  Show the field ONLY if:  [sr_coi] = '1' | Record verbatim the conflict(s) of interest disclosed by the authors: | Notes |
|  | 43 | sr_affiliat | What type of affiliation did the corresponding author of the SR have? | Radio 1 \| Food industry; 4 \| Other industry; 2 \| Non-industry; 3 \| Mixed; 5 \| Unclear |
|  | 44 | ma_n_studies | Section Header: *General characteristics of the index meta-analysis (i.e. [index_ma])*  Specify the total number of studies included in the index meta-analysis (stated on page 1 of PDF): | Text |
|  | 45 | ma_n_patients | Specify the total number of participants included in the index meta-analysis (sum values presented on the forest plot/table or type 'NR' if sample sizes were not available): | Text |
|  | 46 | ma_patients | Specify the type of participants included in studies included in the index meta-analysis (e.g. "generally healthy", "normal weight"): | Text |
|  | 47 | ma_intervention | Specify the intervention/exposure investigated in the index meta-analysis (e.g. "red meat 3 times a week"): | Text |
|  | 48 | ma_comparator | Specify the comparator investigated in the index meta-analysis (e.g. "red meat once a month"): | Text |
|  | 49 | ma_study_type | What type of studies were included in the index meta-analysis? | Radio 1 \| Randomized trial; 2 \| Non-randomized study; 3 \| Both randomized trials and non-randomized studies |
|  | 50 | ma_study_type_spec | Indicate the designs of the studies included in the index meta-analysis as specified by the systematic reviewers (e.g. cohort study, case-control study): | Text |
|  | 51 | ma_outcome_spec | Record how the index meta-analysis was specified (e.g. "weight" or "weight at 6 weeks" or "change in weight at 6 weeks"): | Text |
|  | 52 | ma_domain | Specify the outcome domain investigated in the index meta-analysis (e.g. all-cause mortality, body weight): | Text |
|  | 53 | ma_outcome_label | Specify the label given by the review authors to the index meta-analysis outcome: | Radio 1 \| Primary; 2 \| Secondary; 3 \| Unlabelled |
|  | 54 | ma_model | Specify the model for the index meta-analysis: | Radio 1 \| Fixed-effect; 2 \| Random-effects; 3 \| Unclear |
|  | 55 | sr_e | Section Header: *Eligibility criteria and decision rules*  Review authors reported...(select ALL of the following that apply): | Checkbox 1 \| sr_e___1 \| ...eligible measurement instruments for [index_ma]; 2 \| sr_e___2 \| ...eligible definitions/diagnostic criteria for [index_ma]; 3 \| sr_e___3 \| ...eligible cut-points on an instrument for [index_ma]; 4 \| sr_e___4 \| ...eligible time points for [index_ma]; 5 \| sr_e___5 \| ...eligible interventions/exposures; 6 \| sr_e___6 \| ...whether final values or change from baseline values (or both) for [index_ma] were eligible; 7 \| sr_e___7 \| ...which analysis samples for [index_ma] were eligible (e.g. intention-to-treat, per-protocol or as-treated); 8 \| sr_e___8 \| ...whether unadjusted or covariate-adjusted analyses (or both) for [index_ma] were eligible; 9 \| sr_e___9 \| ...whether period or paired analyses (or both) for [index_ma] were eligible (from crossover trials); 10 \| sr_e___10 \| ...which information sources were eligible (e.g. journal article, trials results register) |
|  | 56 | sr_dr | Review authors reported...(select ALL of the following that apply): | Checkbox 1 \| sr_dr___1 \| ...a decision rule to handle results arising from multiple measurement instruments for [index_ma]; 2 \| sr_dr___2 \| ...a decision rule to handle results arising from multiple definitions/diagnostic criteria for [index_ma]; 3 \| sr_dr___3 \| ...a decision rule to handle results arising from multiple cut-points on an instrument for [index_ma]; 4 \| sr_dr___4 \| ...a decision rule to handle results arising from multiple time points for [index_ma]; 5 \| sr_dr___5 \| ...a decision rule to handle results arising from multiple interventions/exposures; 6 \| sr_dr___6 \| ...a decision rule to handle results arising from both final values and change from baseline values for [index_ma]; 7 \| sr_dr___7 \| ...a decision rule to handle results arising from multiple analysis samples (e.g. intention-to-treat, per-protocol or as-treated) for [index_ma]; 8 \| sr_dr___8 \| ...a decision rule to handle results arising from multiple unadjusted or covariate-adjusted analyses for [index_ma]; 9 \| sr_dr___9 \| ...a decision rule to handle results arising from period and paired analyses for [index_ma] in crossover trials; 10 \| sr_dr___10 \| ...a decision rule to handle results arising from multiple information sources (e.g. journal article, trials results register); 11 \| sr_dr___11 \| ...a decision rule to handle results arising from overlapping samples of participants (e.g. subgroup of men of all ages and subgroup of older adults which includes men); 12 \| sr_dr___12 \| ...a decision rule to handle results arising from another source of multiplicity |
|  | 57 | sr_e_scale_txt  Show the field ONLY if:  [sr_e(1)] = '1' | Specify the eligible measurement instruments for [index_ma]: | Notes |
|  | 58 | sr_dr_scale_txt  Show the field ONLY if:  [sr_dr(1)] = '1' | Specify the decision rule to handle results arising from multiple measurement instruments for [index_ma]: | Notes |
|  | 59 | sr_e_event_txt  Show the field ONLY if:  [sr_e(2)] = '1' | Specify the eligible definitions/diagnostic criteria for [index_ma]: | Notes |
|  | 60 | sr_dr_event_txt  Show the field ONLY if:  [sr_dr(2)] = '1' | Specify the decision rule to handle results arising from multiple definitions/diagnostic criteria for [index_ma]: | Notes |
|  | 61 | sr_e_cutpoint_txt  Show the field ONLY if:  [sr_e(3)] = '1' | Specify the eligible cut-points on an instrument for [index_ma]: | Notes |
|  | 62 | sr_dr_cutpoint_txt  Show the field ONLY if:  [sr_dr(3)] = '1' | Specify the decision rule to handle results arising from multiple cut-points on an instrument for [index_ma]: | Notes |
|  | 63 | sr_e_time_txt  Show the field ONLY if:  [sr_e(4)] = '1' | Specify the eligible time points for [index_ma]: | Notes |
|  | 64 | sr_dr_time_txt  Show the field ONLY if:  [sr_dr(4)] = '1' | Specify the decision rule to handle results arising from multiple time points for [index_ma]: | Notes |
|  | 65 | sr_e_group_txt  Show the field ONLY if:  [sr_e(5)] = '1' | Specify the eligible interventions/exposures (incl. comparators): | Notes |
|  | 66 | sr_dr_group_txt  Show the field ONLY if:  [sr_dr(5)] = '1' | Specify the decision rule to handle results arising from multiple interventions/exposures: | Notes |
|  | 67 | sr_e_fvcs_txt  Show the field ONLY if:  [sr_e(6)] = '1' | Specify which values (i.e. final, change from baseline or both) for [index_ma] were eligible: | Notes |
|  | 68 | sr_dr_fvcs_txt  Show the field ONLY if:  [sr_dr(6)] = '1' | Specify the decision rule to handle results arising from both final and change from baseline values for [index_ma]: | Notes |
|  | 69 | sr_e_itt_txt  Show the field ONLY if:  [sr_e(7)] = '1' | Specify which analysis samples (e.g. intention-to-treat, per-protocol) for [index_ma] were eligible: | Notes |
|  | 70 | sr_dr_itt_txt  Show the field ONLY if:  [sr_dr(7)] = '1' | Specify the decision rule to handle results arising from multiple analysis samples (e.g. intention-to-treat, per-protocol) for [index_ma]: | Notes |
|  | 71 | sr_e_unadj_txt  Show the field ONLY if:  [sr_e(8)] = '1' | Specify which unadjusted and covariate-adjusted analyses for [index_ma] were eligible: | Notes |
|  | 72 | sr_dr_unadj_txt  Show the field ONLY if:  [sr_dr(8)] = '1' | Specify the decision rule to handle results arising from multiple unadjusted and covariate-adjusted analyses for [index_ma]: | Notes |
|  | 73 | sr_e_cross_txt  Show the field ONLY if:  [sr_e(9)] = '1' | Specify which analyses in crossover trials (e.g. first period, second period, paired) were eligible: | Notes |
|  | 74 | sr_dr_cross_txt  Show the field ONLY if:  [sr_dr(9)] = '1' | Specify the decision rule to handle results arising from period and paired analyses in crossover trials: | Notes |
|  | 75 | sr_e_source_txt  Show the field ONLY if:  [sr_e(10)] = '1' | Specify which information sources (e.g. journal article, trials results register) were eligible: | Notes |
|  | 76 | sr_dr_source_txt  Show the field ONLY if:  [sr_dr(10)] = '1' | Specify the decision rule to handle results arising from multiple information sources (e.g. journal article, trials results register): | Notes |
|  | 77 | sr_dr_overlap_txt  Show the field ONLY if:  [sr_dr(11)] = '1' | Specify the decision rule to handle results arising from overlapping samples of participants: | Notes |
|  | 78 | sr_dr_other_txt  Show the field ONLY if:  [sr_dr(12)] = '1' | Specify the decision rule(s) to handle results arising from other sources of multiplicity for [index_ma]: | Notes |
|  | 79 | sr_notes | Notes | Notes |
|  | 80 | sr_characteristics_complete | Section Header: *Form Status*  Complete? | Dropdown 0 \| Incomplete; 1 \| Unverified; 2 \| Complete |
| Instrument: **Meta-analysis study data** (metaanalysis_study_data) | | | | |
|  | 81 | ma_study_id | Study ID (record the ID appearing on the forest plot/table for the index meta-analysis): | Text |
|  | 82 | ma_st_measure | Specify the study effect measure for [index_ma]: | Radio 1 \| Risk ratio; 2 \| Odds ratio; 3 \| Hazard ratio; 4 \| Mean difference; 5 \| Standardized mean difference |
|  | 83 | ma_st_event1  Show the field ONLY if:  [ma_st_measure] = '1' or [ma_st_measure] = '2' or [ma_st_measure] = '3' | Number of events in intervention/exposure group (type 'NR' if not reported): | Text |
|  | 84 | ma_st_total1  Show the field ONLY if:  [ma_st_measure] = '1' or [ma_st_measure] = '2' or [ma_st_measure] = '3' | Sample size of intervention/exposure group (type 'NR' if not reported): | Text |
|  | 85 | ma_st_event2  Show the field ONLY if:  [ma_st_measure] = '1' or [ma_st_measure] = '2' or [ma_st_measure] = '3' | Number of events in comparator group (type 'NR' if not reported): | Text |
|  | 86 | ma_st_total2  Show the field ONLY if:  [ma_st_measure] = '1' or [ma_st_measure] = '2' or [ma_st_measure] = '3' | Sample size of comparator group (type 'NR' if not reported): | Text |
|  | 87 | ma_st_mean1  Show the field ONLY if:  [ma_st_measure] = '4' or [ma_st_measure] = '5' | Mean of intervention/exposure group (type 'NR' if not reported): | Text |
|  | 88 | ma_st_sd1  Show the field ONLY if:  [ma_st_measure] = '4' or [ma_st_measure] = '5' | Standard deviation of intervention/exposure group (type 'NR' if not reported): | Text |
|  | 89 | ma_st_n1  Show the field ONLY if:  [ma_st_measure] = '4' or [ma_st_measure] = '5' | Sample size of intervention/exposure group (type 'NR' if not reported): | Text |
|  | 90 | ma_st_mean2  Show the field ONLY if:  [ma_st_measure] = '4' or [ma_st_measure] = '5' | Mean of comparator group (type 'NR' if not reported): | Text |
|  | 91 | ma_st_sd2  Show the field ONLY if:  [ma_st_measure] = '4' or [ma_st_measure] = '5' | Standard deviation of comparator group (type 'NR' if not reported): | Text |
|  | 92 | ma_st_n2  Show the field ONLY if:  [ma_st_measure] = '4' or [ma_st_measure] = '5' | Sample size of comparator group (type 'NR' if not reported): | Text |
|  | 93 | ma_st_est | Study effect estimate (type 'NR' if not reported): | Text |
|  | 94 | ma_st_seest | Standard error of the study effect estimate (type 'NR' if not reported): | Text |
|  | 95 | ma_st_lciest | Lower 95% confidence interval of the study effect estimate (type 'NR' if not reported): | Text |
|  | 96 | ma_st_uciest | Upper 95% confidence interval of the study effect estimate (type 'NR' if not reported): | Text |
|  | 97 | ma_st_ci_level | If the confidence interval (CI) was not a 95% CI, specify the CI level (e.g. 90% CI, 99% CI); otherwise leave blank | Text |
|  | 98 | ma_st_direction | Specify the direction of the study effect estimate (note: ignore the statistical significance of the effect, just focus on the direction): | Radio 1 \| Favours intervention or exposure; 2 \| Favours comparator; 3 \| Neutral (e.g. RR is somewhere between 0.95 and 1.05 or MD is somewhere between -0.05 and 0.05); 4 \| Unclear |
|  | 99 | ma_st_retrieve | Did the review authors state that data for this particular result were retrieved from study authors? | Yesno 1 \| Yes; 0 \| No |
|  | 100 | ma_st_manip | Did the review authors state that data for this particular result required algebraic transformation to include in meta-analysis (e.g. convert SE to SD)? | Yesno 1 \| Yes; 0 \| No |
|  | 101 | ma_st_transl | Did the review authors state that data for this particular result originated from a report translated into English? | Yesno 1 \| Yes; 0 \| No |
|  | 102 | ma_st_impute | Did the review authors state that data for this particular result was included in the meta-analysis by using a method of imputation? | Yesno 1 \| Yes; 0 \| No |
|  | 103 | ma_st_impute_txt  Show the field ONLY if:  [ma_st_impute] = '1' | Specify the method of imputation: | Text |
|  | 104 | ma_st_notes | Notes | Notes |
|  | 105 | metaanalysis_study_data_complete | Section Header: *Form Status*  Complete? | Dropdown 0 \| Incomplete; 1 \| Unverified; 2 \| Complete |
| Instrument: **Meta-analytic effect** (metaanalytic_effect) | | | | |
|  | 106 | ma_measure | Specify the meta-analytic effect measure for [index_ma]: | Radio 1 \| Risk ratio; 2 \| Odds ratio; 3 \| Hazard ratio; 4 \| Mean difference; 5 \| Standardized mean difference |
|  | 107 | ma_est | Meta-analytic effect estimate: | Text |
|  | 108 | ma_seest | Standard error of the meta-analytic effect estimate (type 'NR' if not reported): | Text |
|  | 109 | ma_lciest | Lower 95% confidence interval of the meta-analytic effect estimate (type 'NR' if not reported): | Text |
|  | 110 | ma_uciest | Upper 95% confidence interval of the meta-analytic effect estimate (type 'NR' if not reported): | Text |
|  | 111 | ma_ci_level | If the confidence interval (CI) was not a 95% CI, specify the CI level (e.g. 90% CI, 99% CI); otherwise leave blank | Text |
|  | 112 | ma_direction | Specify the direction of the meta-analytic effect estimate (note: ignore the statistical significance of the effect, just focus on the direction): | Radio 1 \| Favours intervention or exposure; 2 \| Favours comparator; 3 \| Neutral (e.g. RR is somewhere between 0.95 and 1.05 or MD is somewhere between -0.05 and 0.05); 4 \| Unclear |
|  | 113 | ma_location | Specify the location of the meta-analysis in the article (e.g. Figure 1, Supp Table 2): | Text |
|  | 114 | ma_conclusion | What type of conclusion did the authors draw about the intervention/exposure with regards to [index_ma]? | Radio 1 \| Favourable (e.g. stated that the food/diet had a beneficial/positive effect on [index_ma]); 2 \| Unfavourable (e.g. stated that the food/diet had a harmful/negative effect on [index_ma]); 3 \| Neutral (e.g. stated that the food/diet had neither a beneficial or harmful effect on [index_ma]); 4 \| Unclear; 5 \| No conclusion about [index_ma] drawn |
|  | 115 | ma_conclusion_ab_txt  Show the field ONLY if:  [ma_conclusion] = '1' or [ma_conclusion] = '2' or [ma_conclusion] = '3' or [ma_conclusion] = '5' | Extract verbatim the conclusion reported in the abstract: | Notes |
|  | 116 | ma_conclusion_txt  Show the field ONLY if:  [ma_conclusion] = '1' or [ma_conclusion] = '2' or [ma_conclusion] = '3' or [ma_conclusion] = '4' | Extract verbatim the conclusion reported in the main text: | Notes |
|  | 117 | ma_notes | Notes | Notes |
|  | 118 | metaanalytic_effect_complete | Section Header: *Form Status*  Complete? | Dropdown 0 \| Incomplete; 1 \| Unverified; 2 \| Complete |
| Instrument: **Study data**(study_data) | | | | |
|  | 119 | study_id | Study ID (record the ID in the filename of the PDF): | Text |
|  | 120 | st_data_avail | Was there enough data available in the paper to extract or calculate a result compatible with the index meta-analysis ([index_ma])? | Yesno 1 \| Yes; 0 \| No |
|  | 121 | st_design  Show the field ONLY if:  [st_data_avail] = '1' | Indicate the design of the study (e.g. parallel-group randomized trial, crossover trial, cohort study, case-control study): | Text |
|  | 122 | st_measure  Show the field ONLY if:  [st_data_avail] = '1' | Specify the study effect measure for a result compatible with the index meta-analysis ([index_ma]): | Radio 0 \| Risk ratio; 1 \| Odds ratio; 2 \| Hazard ratio; 3 \| Rate ratio; 4 \| Prevalence ratio; 5 \| Mean difference; 6 \| Standardized mean difference |
|  | 123 | st_def_event  Show the field ONLY if:  [st_measure] = '0' or [st_measure] = '1' or [st_measure] = '2' or [st_measure] = '3' or [st_measure] = '4' | Specify the definition/diagnostic criteria for the event (type 'NR' if not reported): | Text |
|  | 124 | st_scale  Show the field ONLY if:  [st_measure] = '5' or [st_measure] = '6' | Specify the outcome measurement instrument used to generate the result (type 'NR' if not reported): | Text |
|  | 125 | st_intervention  Show the field ONLY if:  [st_data_avail] = '1' | Specify the intervention/exposure (e.g. 'red meat 3 times a week'): | Text |
|  | 126 | st_comparator  Show the field ONLY if:  [st_data_avail] = '1' | Specify the comparator (e.g. 'red meat once a month'): | Text |
|  | 127 | st_time  Show the field ONLY if:  [st_data_avail] = '1' | Specify the time point for the result (e.g. 6 weeks; type 'NR' if not reported): | Text |
|  | 128 | st_itt  Show the field ONLY if:  [st_data_avail] = '1' | Specify the analysis sample for the result (type 'NR' if not reported): | Radio 1 \| Intention-to-treat (ITT); 2 \| Modified ITT; 3 \| Per-protocol; 4 \| As-treated; 5 \| Other; 6 \| Unclear |
|  | 129 | st_unadj  Show the field ONLY if:  [st_data_avail] = '1' | Was the result unadjusted or adjusted for covariates? | Radio 1 \| Unadjusted; 2 \| Adjusted; 3 \| Unclear |
|  | 130 | st_covar  Show the field ONLY if:  [st_unadj] = '2' | Which covariates were included in the model? | Text |
|  | 131 | st_other_id  Show the field ONLY if:  [st_data_avail] = '1' | Specify any other identifying characteristic of the result, e.g. 'subgroup analysis of men only' (type 'NA' if not applicable): | Text |
|  | 132 | st_event1  Show the field ONLY if:  [st_measure] = '0' or [st_measure] = '1' or [st_measure] = '2' or [st_measure] = '3' or [st_measure] = '4' | Number of events in intervention/exposure group (type 'NR' if not reported): | Text |
|  | 133 | st_total1  Show the field ONLY if:  [st_measure] = '0' or [st_measure] = '1' or [st_measure] = '2' or [st_measure] = '3' or [st_measure] = '4' | Sample size of intervention/exposure group (type 'NR' if not reported): | Text |
|  | 134 | st_event2  Show the field ONLY if:  [st_measure] = '0' or [st_measure] = '1' or [st_measure] = '2' or [st_measure] = '3' or [st_measure] = '4' | Number of events in comparator group (type 'NR' if not reported): | Text |
|  | 135 | st_total2  Show the field ONLY if:  [st_measure] = '0' or [st_measure] = '1' or [st_measure] = '2' or [st_measure] = '3' or [st_measure] = '4' | Sample size of comparator group (type 'NR' if not reported): | Text |
|  | 136 | st_fvcs  Show the field ONLY if:  [st_measure] = '5' or [st_measure] = '6' | Which values were used to calculate means? | Radio 1 \| Final values; 2 \| Change from baseline values; 3 \| Unclear |
|  | 137 | st_meansd  Show the field ONLY if:  [st_measure] = '5' or [st_measure] = '6' | Were means and standard deviations reported for each group? | Yesno 1 \| Yes; 0 \| No |
|  | 138 | st_mean1  Show the field ONLY if:  [st_measure] = '5' or [st_measure] = '6' | Mean of intervention/exposure group (type 'NR' if not reported): | Text |
|  | 139 | st_sd1  Show the field ONLY if:  [st_measure] = '5' or [st_measure] = '6' | Standard deviation of intervention/exposure group (type 'NR' if not reported): | Text |
|  | 140 | st_n1  Show the field ONLY if:  [st_measure] = '5' or [st_measure] = '6' | Sample size of intervention/exposure group (type 'NR' if not reported or 'Paired N' if this was a before-after comparison): | Text |
|  | 141 | st_mean2  Show the field ONLY if:  [st_measure] = '5' or [st_measure] = '6' | Mean of comparator group (type 'NR' if not reported): | Text |
|  | 142 | st_sd2  Show the field ONLY if:  [st_measure] = '5' or [st_measure] = '6' | Standard deviation of comparator group (type 'NR' if not reported): | Text |
|  | 143 | st_n2  Show the field ONLY if:  [st_measure] = '5' or [st_measure] = '6' | Sample size of comparator group (type 'NR' if not reported or 'Paired N' if this was a before-after comparison): | Text |
|  | 144 | st_semean1  Show the field ONLY if:  [st_meansd] = '0' | Standard error of the mean of the intervention/exposure group (type 'NR' if not reported): | Text |
|  | 145 | st_lcimean1  Show the field ONLY if:  [st_meansd] = '0' | Lower 95% confidence interval of the mean of the intervention/exposure group (type 'NR' if not reported): | Text |
|  | 146 | st_ucimean1  Show the field ONLY if:  [st_meansd] = '0' | Upper 95% confidence interval of the mean of the intervention/exposure group (type 'NR' if not reported): | Text |
|  | 147 | st_semean2  Show the field ONLY if:  [st_meansd] = '0' | Standard error of the mean of the comparator group (type 'NR' if not reported): | Text |
|  | 148 | st_lcimean2  Show the field ONLY if:  [st_meansd] = '0' | Lower 95% confidence interval of the mean of the comparator group (type 'NR' if not reported): | Text |
|  | 149 | st_ucimean2  Show the field ONLY if:  [st_meansd] = '0' | Upper 95% confidence interval of the mean of the comparator group (type 'NR' if not reported): | Text |
|  | 150 | st_est  Show the field ONLY if:  [st_data_avail] = '1' | Study effect estimate (type 'NR' if not reported): | Text |
|  | 151 | st_seest  Show the field ONLY if:  [st_data_avail] = '1' | Standard error of the study effect estimate (type 'NR' if not reported): | Text |
|  | 152 | st_lciest  Show the field ONLY if:  [st_data_avail] = '1' | Lower 95% confidence interval of the study effect estimate (type 'NR' if not reported): | Text |
|  | 153 | st_uciest  Show the field ONLY if:  [st_data_avail] = '1' | Upper 95% confidence interval of the study effect estimate (type 'NR' if not reported): | Text |
|  | 154 | st_ci_level | If the confidence interval (CI) was not a 95% CI, specify the CI level (e.g. 90% CI, 99% CI); otherwise leave blank | Text |
|  | 155 | st_pvalue  Show the field ONLY if:  [st_data_avail] = '1' | Specify the exact P-value for the study effect estimate (type 'NR' if not reported): | Text |
|  | 156 | st_direction  Show the field ONLY if:  [st_data_avail] = '1' | Specify the direction of the study effect estimate (note: ignore the statistical significance of the effect, just focus on the direction): | Radio 1 \| Favours intervention or exposure; 2 \| Favours comparator; 3 \| Neutral (e.g. RR is somewhere between 0.95 and 1.05 or MD is somewhere between -0.05 and 0.05); 4 \| Unclear |
|  | 157 | st_location  Show the field ONLY if:  [st_data_avail] = '1' | Specify the location of the data in the report (e.g. Table 2, Figure 1, text page 378): | Text |
|  | 158 | incl_ma  Show the field ONLY if:  [st_data_avail] = '1' | Does the study effect estimate and 95% CI match the one included in the index meta-analysis? (note: if the study effect estimate and 95% CI were not reported in the study paper, calculate these values from the summary statistics) | Radio 1 \| Yes; 2 \| No; 3 \| Unclear |
|  | 159 | st_error  Show the field ONLY if:  [incl_ma] = '1' | Were the data entered by the systematic reviewers incorrect in any way (e.g. wrong sample size for cross-over trials, or standard error entered as standard deviation)? | Yesno 1 \| Yes; 0 \| No |
|  | 160 | st_error_text  Show the field ONLY if:  [st_error] = '1' | Describe how the data were entered incorrectly by the systematic reviewers: | Notes |
|  | 161 | st_notes | Notes | Notes |
|  | 162 | study_data_complete | Section Header: *Form Status*  Complete? | Dropdown 0 \| Incomplete; 1 \| Unverified; 2 \| Complete |

**Supplementary Table S4: Data sources and data items (reproduced from Kanukula et al. J Clin Epidemiol 2022;142:171-183).**

| **Source** | **Data items** |
| --- | --- |
| Systematic review protocol | Year of publication/registration; eligibility criteria and decision rules to select results to include in the index meta-analysis |
| Systematic review | *General characteristics of the systematic review* |
|  | Journal name; year of publication; corresponding author’s country and affiliation; conflicts of interest of review authors; source of funding for the review; |
|  | *General characteristics of the index meta-analysis* |
|  | Number of studies and participants; type of population investigated; type of interventions/exposures investigated; type of studies included in the meta-analysis; outcome domain (such as weight, cardiovascular function); outcome primacy label (primary or secondary or unlabelled); meta-analysis effect measure; meta-analysis model; eligibility criteria and decision rules to select results to include in the index meta-analysis; summary statistics, effect estimates and measures of precision (e.g confidence interval) for each included study; and the meta-analytic effect estimate and measure of precision. |
| Study reports | *Outcome data that could potentially be included in the index meta-analysis* |
|  | Outcome definition and measurement instrument; intervention/exposure description; comparator description; time point; analysis sample (e.g. intention-to-treat, per-protocol); summary statistics (e.g. number of events and sample sizes of both intervention/exposure and comparator); effect measure (e.g. risk ratio, mean difference); effect estimates and measures of precision (e.g. 95% confidence interval) and location of data in the report; whether results were unadjusted or adjusted for covariates, covariates that were adjusted for (if applicable). |

**Supplementary Table S5. Mathematical details of the construction of the Potential Bias Index (PBI) and associated statistical test (reproduced from Page et al. Syst Rev 2013;2:21).**

Here we provide the mathematical details of a summary statistic which we call the Potential Bias Index (PBI). In brief, PBI measures the average location of the rank of the selected effect estimates on a scale from 0 to 1, where 0 represents the lowest possible rank and 1 represents highest possible rank, and 0.5 represents the middle value. Consider a situation with *k* trials, labeled i=1,2, …, k, and for the *i’th* trial there are *n_i_* effect estimates. Order these effect estimates from smallest to largest, with the smallest getting rank 1 and the largest rank n_i_. Let X_i_ denote the rank of the effect estimate that was actually chosen for reporting in trial i. Rescale X_i_ so that it can take values between 0 and 1, with the value 0 when X_i_ is the lowest rank and the value 1 when X_i_ is the highest rank. Call this variable Y_i_ and define it as $Y_{i}=\frac{X_{i}-1}{n_{i}-1}$. We define the PBI as a weighted average of the Y_i_’s , with weights equal to the number of effect measures in each trial n*i*,

$$PBI=\sum_{i=1}^{k} n_{i}Y_{i} /\sum_{i=1}^{k} n_{i}=\sum_{i=1}^{k} \frac{n_{i}\left( X_{i}-1 \right)}{n_{i}-1} /\sum_{i=1}^{k} n_{i}$$

Under an assumption of random selection, X_i_ could be any one of the effect estimates with equal probability and therefore $P\left( X_{i}=j \right)=1/n_{i}$ for each j=1,2,…,n_i_. It is then straightforward to calculate that the expected value of X_i_ under randomness is $\frac{n_{i}}{2}+\frac{1}{2}$, the expected value of Y_i_ is 0.5, and therefore E[PBI] = 0.5$E\left( T \right)=\frac{1}{2}+\frac{1}{2}n^{*}$. Similarly, the variance of PBI can be calculated as

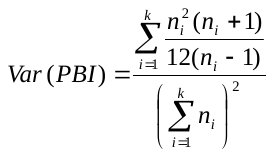

A statistical test of the null hypothesis of randomness can be constructed using $Z=(PBI-E\left( PBI \right))/\sqrt{Var(PBI)}$ which should follow a standard normal distribution under the hypothesis of randomness when the number of trials is sufficiently large.

We assessed the properties of this statistical test under the assumption of randomness by simulating a set of trials, each with a number of effect estimates, and then randomly selecting one of the effect estimates with equal probability and comparing its ranking with the other effect sizes. Specifically, we generated between k=3 and k=120 trials, with the number of effect estimates per trial chosen from a truncated Poisson distribution of mean 3 or 6, where the truncation excluded values of zero or one effect estimate so that each trial contained two or more effect estimates. The reported effect estimate was then randomly chosen from the set of effect estimates for each trial. The statistic PBI was then calculated for each set of trials, and assessment of its statistical significance at the 5% level made by conversion to the test statistic Z as above and comparison with the standard normal distribution. Over 20,000 replications of each configuration, the percentage of Z statistics where the hypothesis of randomness was rejected was recorded, i.e. the Type I error rate. The simulation standard error for a nominal 5% Type I error rate with 20,000 replications was +/- 0.15%:

| Number of trials | Average number of effect measures per trial | Empirical Type I error rate % |
| --- | --- | --- |
| 3 | 3 | 4.86 |
| 3 | 6 | 4.61 |
| 5 | 3 | 4.58 |
| 5 | 6 | 4.83 |
| 10 | 3 | 5.08 |
| 10 | 6 | 5.11 |
| 15 | 3 | 4.83 |
| 15 | 6 | 5.01 |
| 30 | 3 | 5.03 |
| 30 | 6 | 4.92 |
| 60 | 3 | 5.04 |
| 60 | 6 | 4.92 |
| 120 | 3 | 4.81 |
| 120 | 6 | 5.14 |

The results indicate that the nominal Type I error of 5% is well preserved using this test even for assessments with as few as 3 trials.

*Technical Note: Our definition of* $Y_{i}=\frac{X_{i}-1}{n_{i}-1}$ *in some disciplines (e.g. education, social science, and spread sheet software) is occasionally called the “percentile ranking”, however percentiles have a slightly different definition in the statistical and health sciences and therefore we have refrained from providing any interpretation of Y or PBI in terms of such “percentile ranks”.*

**Supplementary Table S6. Worked example of the Potential Bias Index (reproduced from Page et al. Syst Rev 2013;2:21).**

Consider a review consisting of 10 trials. The number of effect estimates for each trial is provided as well as the rank of the effect estimate chosen among those available for each trial. Suppose the data are the following:

| **Trial** | **Number of effect estimates (n)** | **Rank of chosen effect estimate (X)** | **Location of chosen effect estimate (Y)** |
| --- | --- | --- | --- |
| **1** | 3 | 2 | 0.50 |
| **2** | 7 | 6 | 0.83 |
| **3** | 4 | 3 | 0.67 |
| **4** | 6 | 5 | 0.80 |
| **5** | 4 | 2 | 0.33 |
| **6** | 5 | 4 | 0.75 |
| **7** | 6 | 6 | 1 |
| **8** | 2 | 1 | 0 |
| **9** | 4 | 4 | 1 |
| **10** | 5 | 4 | 0.75 |

For Trial #1, there were 3 effect estimates and the rank of the chosen effect estimate was 2, that is the middle or median value, and its location is therefore halfway between the lowest and highest rank. For Trial #2 there were 7 effect estimates and the chosen estimate had rank 6. There are a total of 6 units of rank between 1 and 7 (i.e. 1 to 2, 2 to 3, 3 to 4, 4 to 5, 5 to 6 and 6 to 7) and the chosen rank of 6 is therefore 5/6ths = 83% of the distance between lowest and highest rank. In general, the rank location Y is calculated as (X-1)/(n-1).

The statistic PBI =
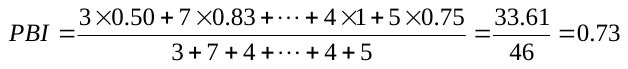

Therefore on average the effect estimates chosen were 73% of the distance between the smallest and largest rank, i.e. approximately halfway between the middle rank and the maximum.

The standard error of the PBI can be calculated to be 0.118, and therefore the Z-statistic equals (0.73-0.50)/0.118 = 1.97, with a two-tailed p-value of 0.049. This indicates some evidence that the effect estimate selection is systematically higher than that expected by random selection.

A 95% confidence interval for the PBI is obtained from 1000 bootstrap replications as 0.58 to 0.85.

**Supplementary Table S7. Case study of the impact of selection of study effect estimates on meta-analyses (reproduced from Page et al. F1000Research 2019;8:1760).**

In our previous study of selective inclusion of results (PMID 27121706), we included a meta-analysis conducted by Baumeister et al., which included two trials examining the effect of psychological interventions versus usual care for depression (PMID 21901717). Study 1 had three effect estimates for depression that were compatible with the meta-analysis (1A, 1B and 1C) and Study 2 had two effect estimates for depression that were compatible with the meta-analysis (2A and 2B). There were six meta-analyses that could be generated based on the data available in these two studies (see possible combinations in Table 1, and results of each meta-analysis in Table 2).

**Table 1. Effect estimates in each of the six meta-analyses that could be generated from the two studies included in the review by Baumeister et al.**

|  | Index meta-analysis | Meta-analysis 2 | Meta-analysis 3 | Meta-analysis 4 | Meta-analysis 5 | Meta-analysis 6 |
| --- | --- | --- | --- | --- | --- | --- |
| Study 1 | 1A | 1A | 1B | 1B | 1C | 1C |
| Study 2 | 2A | 2B | 2A | 2B | 2A | 2B |

**Table 2. Results of the six random-effects meta-analyses of standardised mean differences (SMD) that could be generated from the two studies included in the review by Baumeister et al.**

|  | Index meta-analysis | Meta-analysis 2 | Meta-analysis 3 | Meta-analysis 4 | Meta-analysis 5 | Meta-analysis 6 |
| --- | --- | --- | --- | --- | --- | --- |
| Study 1 | 0.12 (-0.46, 0.70) | 0.12 (-0.46, 0.70) | -0.38 (-0.95, 0.19) | -0.38 (-0.95, 0.19) | -0.13 (-0.69, 0.42) | -0.13 (-0.69, 0.42) |
| Study 2 | -0.81 (-1.26, -0.36) | -0.97 (-1.43, -0.51) | -0.81 (-1.26, -0.36) | -0.97 (-1.43, -0.51) | -0.81 (-1.26, -0.36) | -0.97 (-1.43, -0.51) |
| Meta-analysis | -0.36 (-1.27, 0.55) | -0.44 (-1.51, -0.63) | -0.63 (-1.05, -0.22) | -0.70 (-1.28, -0.12) | -0.49 (-1.16, 0.18) | -0.57 (-1.39. 0.26) |

Data in each cell are standardised mean difference (95% confidence interval)

The index meta-analytic SMD was -0.364, and the median meta-analytic SMD (across all six meta-analyses) was -0.528. The difference (95% CI) between the index and median meta-analytic SMD was 0.16 (-0.75, 1.07), which suggests that there was no evidence that the index meta-analytic SMD differed to what we would expect to see in a meta-analysis generated when there was no selective inclusion.

**Supplementary Table S8. Number (%) of SR protocols and SRs reporting eligibility criteria and decision rules to select effect estimates to include in meta-analyses**

| **Types of eligibility criterion and decision rule** | **SR protocols**  **n (%)**  **N=13** | **SRs**  **n (%)**  **N=39** |
| --- | --- | --- |
| Total |  |  |
| At least one eligibility criterion | 13 (100) | 39 (100) |
| At least one decision rule | 3 (23) | 27 (69) |
| Measurement instruments |  |  |
| Eligibility criteria | 1 (8) | 1 (3) |
| Decision rule | 0 | 2 (5) |
| Definitions/diagnostic criteria |  |  |
| Eligibility criteria | 0 | 2 (5) |
| Decision rule | 0 | 1 (3) |
| Cut-points on a measurement instrument |  |  |
| Eligibility criteria | 0 | 0 |
| Decision rule | 0 | 0 |
| Time points |  |  |
| Eligibility criteria | 4 (31) | 3 (8) |
| Decision rule | 0 | 3 (8) |
| Interventions/exposures |  |  |
| Eligibility criteria | 12 (92) | 37 (95) |
| Decision rule | 2 (15) | 15 (38) |
| Information sources |  |  |
| Eligibility criteria | 3 (23) | 3 (8) |
| Decision rule | 0 | 3 (8) |
| Analyses |  |  |
| Eligibility criteria for any type of analysis | 2 (15) | 10 (26) |
| Decision rule for any type of analysis | 1 (8) | 14 (36) |
| Rule for final vs. change from baseline values | 0 | 3 (8) |
| Rule for analyses undertaken on multiple samples (e.g., ITT vs. per-protocol) | 0 | 0 |
| Rule for unadjusted vs. covariate-adjusted analyses | 0 | 9 (23) |
| Rule for period vs. paired analyses in crossover randomized trials | 0 | 1 (3) |
| Rule to handle results arising from overlapping samples of participants | 1 (8) | 1 (3) |
| Other decision rule | 0 | 5 (13) |

**Supplementary Table S9: Pre-specified sensitivity analyses for the Potential Bias Index (PBI)**

| **PBI analyses** | **Number of comparisons** | **Number of meta-analyses** | **PBI (95% CI*)** |
| --- | --- | --- | --- |
| **Primary analysis** | 206 | 38 | 0.49 (0.42-0.55) |
| **Sensitivity analyses** |  |  |  |
| 1. Inclusion of the set of study effect estimates that were compatible with the eligibility criteria and decision rules in the methods section of the review | 157 | 33 | 0.49 (0.42-0.57) |
| 2. Inclusion of study effect estimates that required algebraic manipulation | 224 | 39 | 0.48 (0.43-0.54) |
| 3. Exclusion of study effect estimates in meta-analyses comparing one type of food/diet with another | 150 | 23 | 0.48 (0.41-0.55) |

*Percentile-based confidence intervals for the PBI were constructed using bootstrap methods by resampling individual trials 2,000 times.

**Supplementary Table S10: Peer-reviewer suggested sensitivity analysis for the Potential Bias Index (PBI)**

| **PBI analyses** | **No. of comparisons** | **Number of meta-analyses** | **PBI (95% CI)** | ***I^2^*** | **p-value for heterogeneity** | ${\hat{\boldsymbol{\tau}}}^{\boldsymbol{2}}\boldsymbol{(}\boldsymbol{REML}\boldsymbol{)}$ **(95% CI)** | **Prediction interval (95%)** |
| --- | --- | --- | --- | --- | --- | --- | --- |
| **Primary analysis** | 206 | 38 | 0.49 (0.42-0.55)^a^ |  | Not applicable | Not applicable | Not applicable |
| **Sensitivity analyses** |  |  |  |  |  |  |  |
| 1. Random-effects meta-analysis of meta-analysis PBI estimates (see Supplementary Figure S10 for forest plot) | 206 | 38 | 0.49 (0.43-0.55) | 0% | 0.641 | 0.000 (0.000-0.016) | (0.43-0.55) |
| 2. Random-effects meta-analysis of meta-analysis PBI estimates. Restricted to meta-analyses with at least 3 studies^b^ | 187 | 25 | 0.48 (0.42-0.55) | 0% | 0.497 | 0.000 (0.000-0.018) | (0.42-0.55) |
| 3. Random-effects meta-analysis of meta-analysis PBI estimates. Restricted to meta-analyses where the confidence limits for the meta-analysis PBI estimates both fell within the range 0 and 1^b^ | 150 | 16 | 0.49 (0.41-0.56) | 0% | 0.874 | 0.000 (0.000-0.016) | (0.41-0.56) |

^a^Percentile-based confidence intervals for the PBI were constructed using bootstrap methods by resampling individual trials 2,000 times.

^b^The statistical test associated with the PBI assumes the number of trials is sufficiently large. However, our simulation study established its validity even for a small number of studies (as few as three) under the null hypothesis (Supplementary Table S5). This suggests that variance of the PBI may be accurate even for a small number of studies per meta-analysis. We undertook two additional sensitivity analyses to examine the impact of restricting the included meta-analyses to those which had at least 3 studies (analysis 2 in the table), and those where the confidence limits for the estimated PBI in a particular meta-analysis both fell between 0 and 1. Results from these additional sensitivity analyses did not differ compared with our primary sensitivity analysis (analysis 1 in the table).

Abbreviations: REML, restricted maximum likelihood estimator.

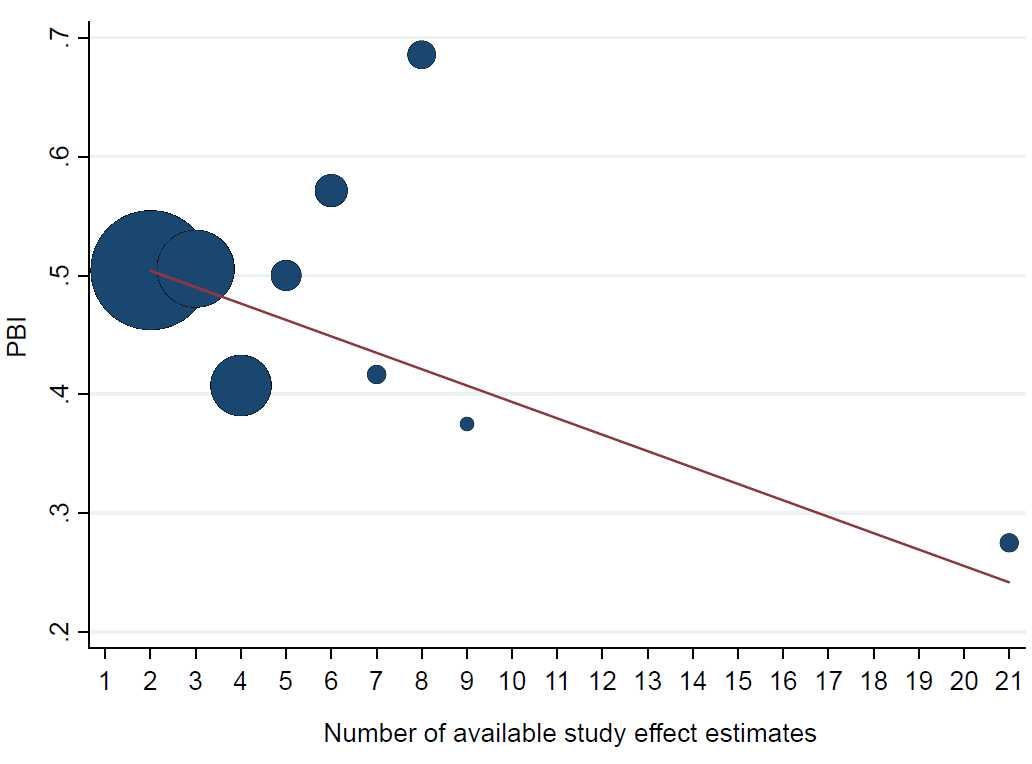

**Supplementary Figure S1. Scatterplot of the relationship between the Potential Bias Index (PBI) and the number of study effect estimates available for inclusion in a meta-analysis. The observed PBI values are depicted by blue dots, the sizes of which are proportional to the number (n) of studies available. The red line represents the fitted regression line, weighted by the number of observations available per data point.**

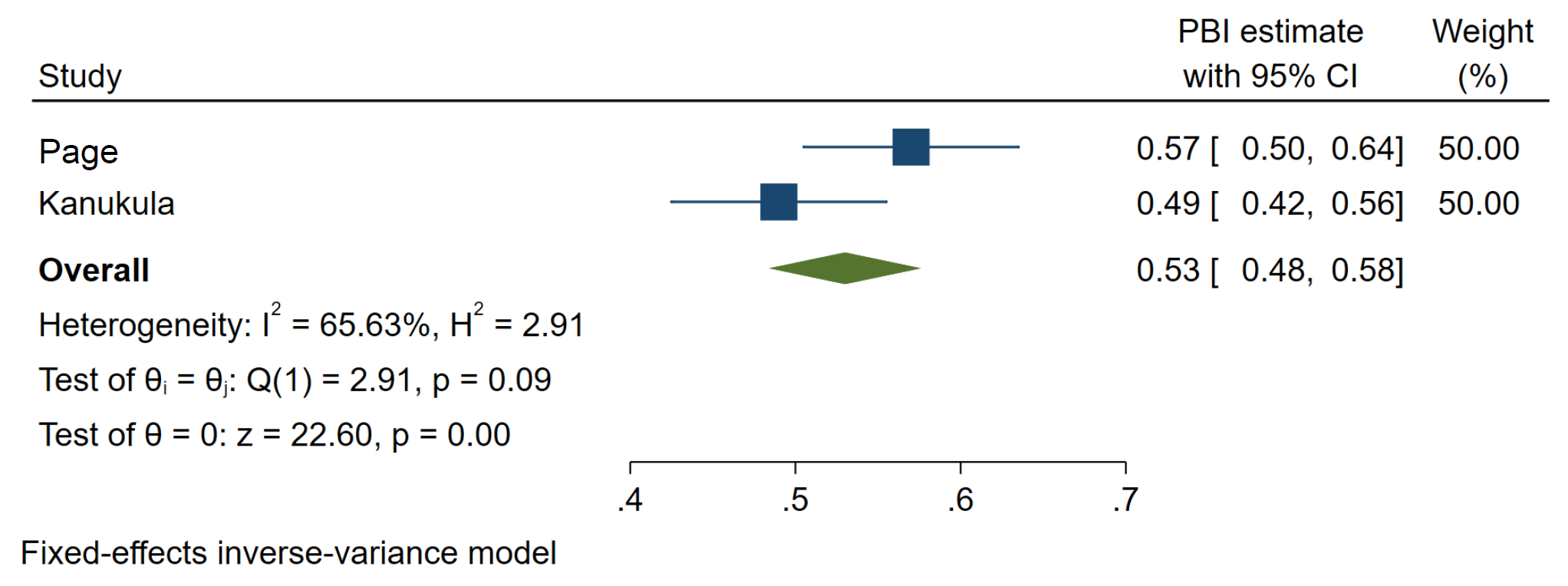

**Supplementary Figure S2. Fixed-effect meta-analysis of Potential Bias Index (PBI) estimates from the current study and a previous study exploring selective inclusion of results. A PBI of 0.5 suggests that there is no evidence of selective inclusion of the most or least favourable effect estimates.**

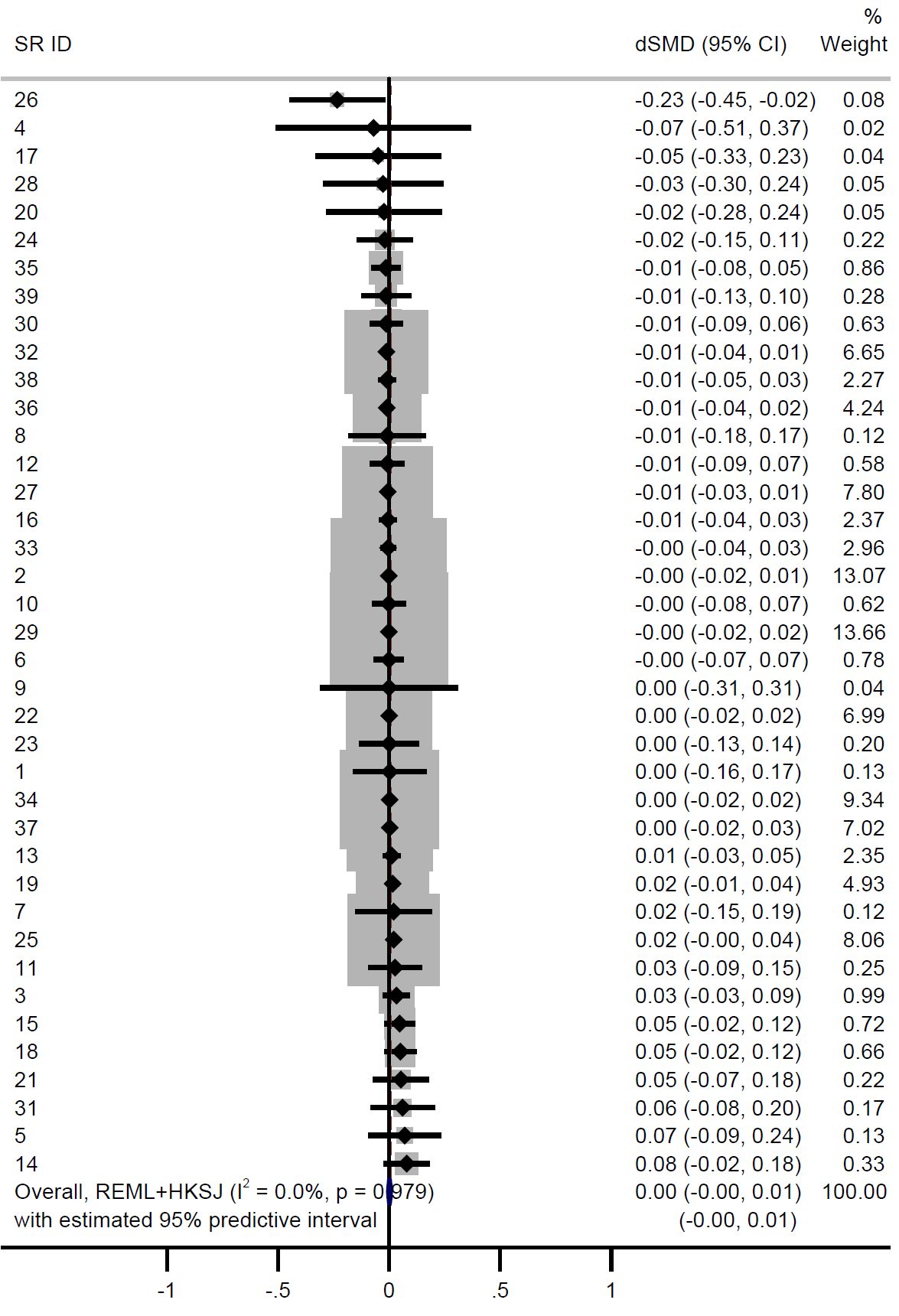

**Supplementary Figure S3. Meta-analysis of differences between the index meta-analytic standardised mean difference (SMD) and median of all its possible meta-analytic SMDs (each calculated using the fixed-effect model). Differences less than zero indicate that the index meta-analytic SMD is more favourable to the intervention compared with the median meta-analytic SMD. dSMD = difference in standardised mean difference.**

When all possible meta-analytic SMDs were calculated using a fixed-effect model, the median of the differences between the index meta-analysis and the median of all its possible meta-analytic SMDs was -0.001 standard deviation units (IQR -0.01 to 0.02; range -0.23 to 0.08). Meta-analysing these differences using a random-effects model yielded a pooled difference of 0.002 standard deviation units (95% CI -0.003 to 0.01; I^2^ = 0%).

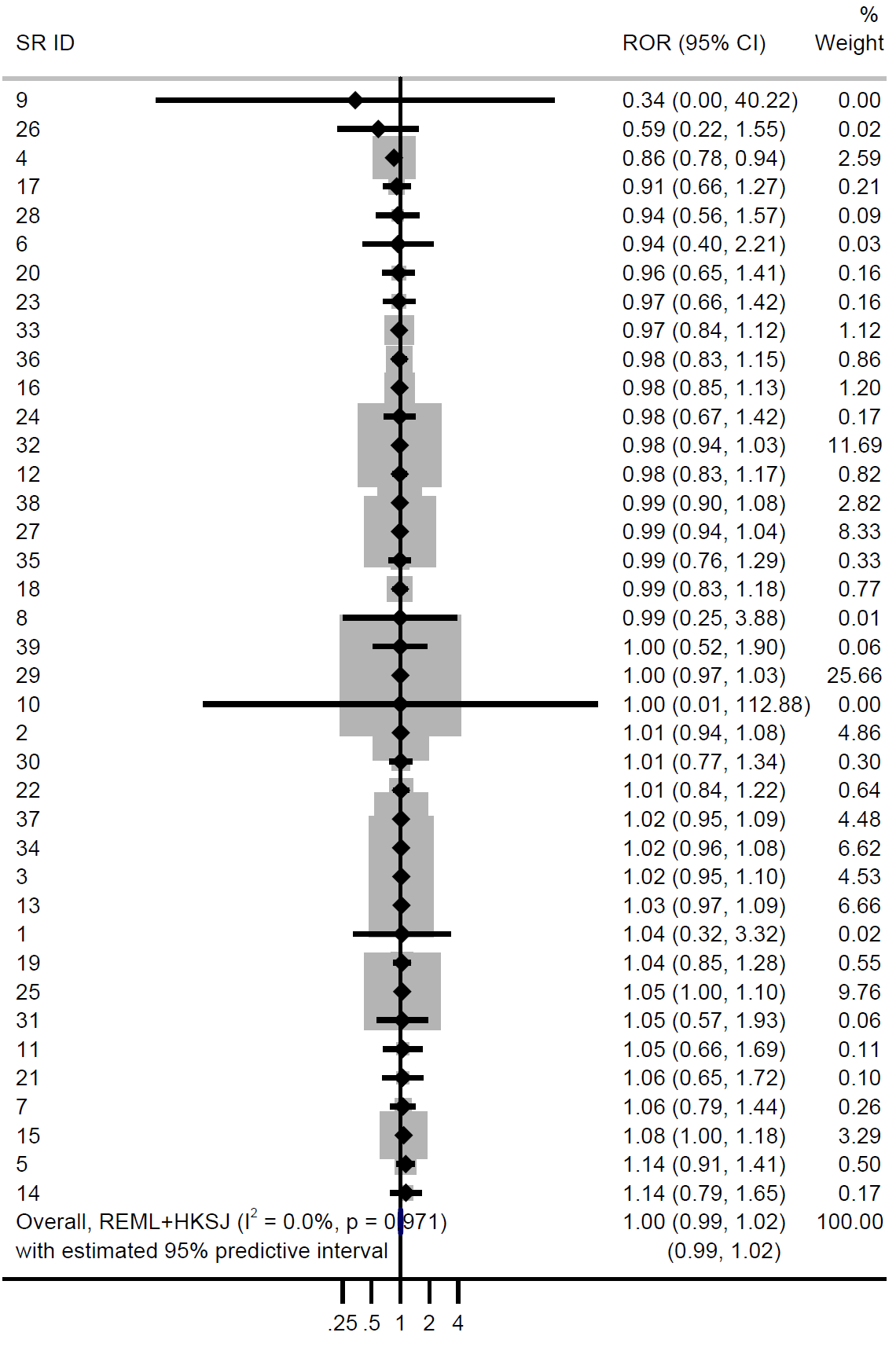

**Supplementary Figure S4. Meta-analysis of differences between the index meta-analytic odds ratio (OR) and median of all its possible meta-analytic ORs (each calculated using the random-effects model). Differences less than 1 indicate that the index meta-analytic OR is more favourable to the intervention compared with the median meta-analytic OR. ROR = ratio of odds ratios.**

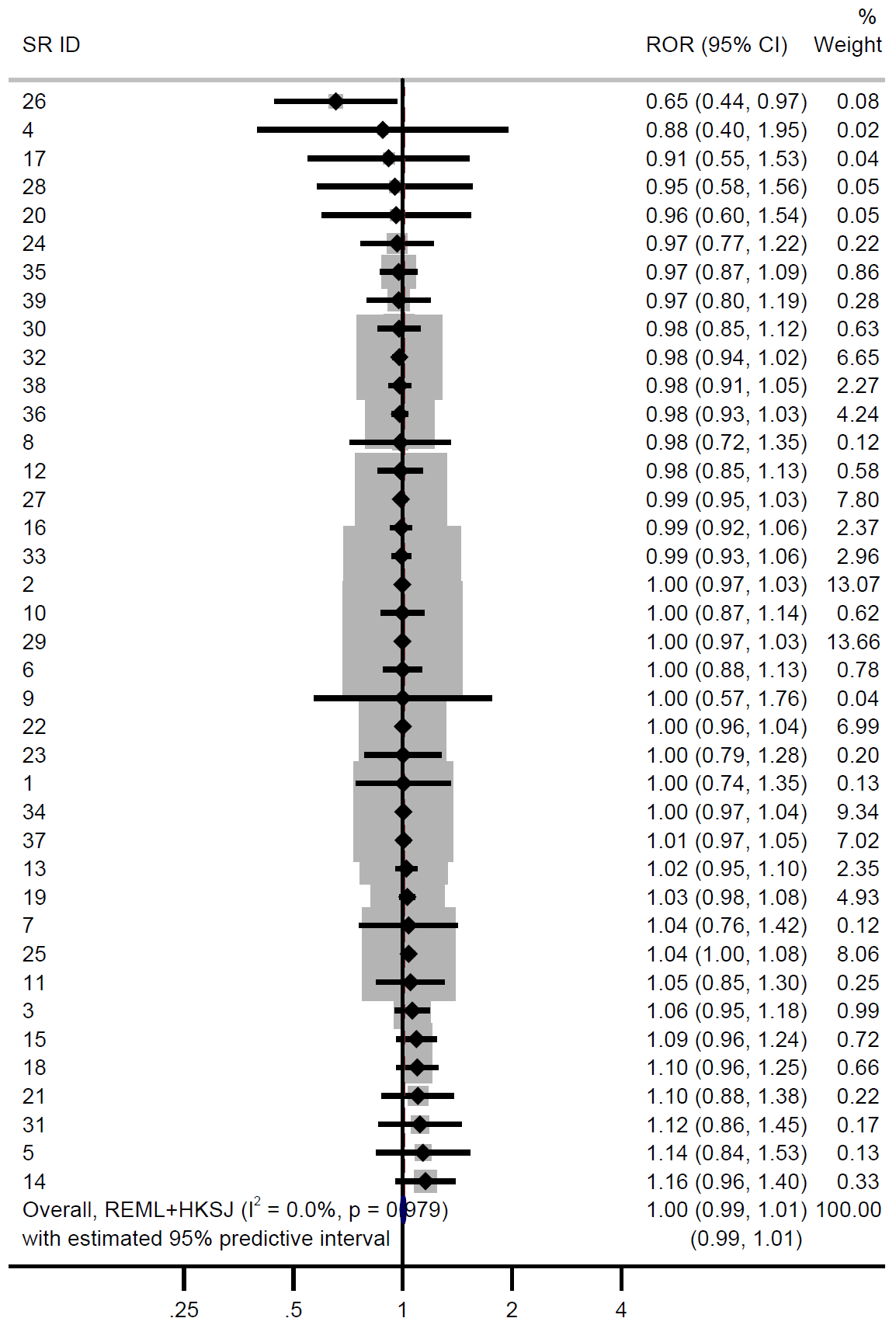

**Supplementary Figure S5. Meta-analysis of differences between the index meta-analytic odds ratio (OR) and median of all its possible meta-analytic ORs (each calculated using the fixed-effect model). Differences less than 1 indicate that the index meta-analytic OR is more favourable to the intervention compared with the median meta-analytic OR. ROR = ratio of odds ratios.**

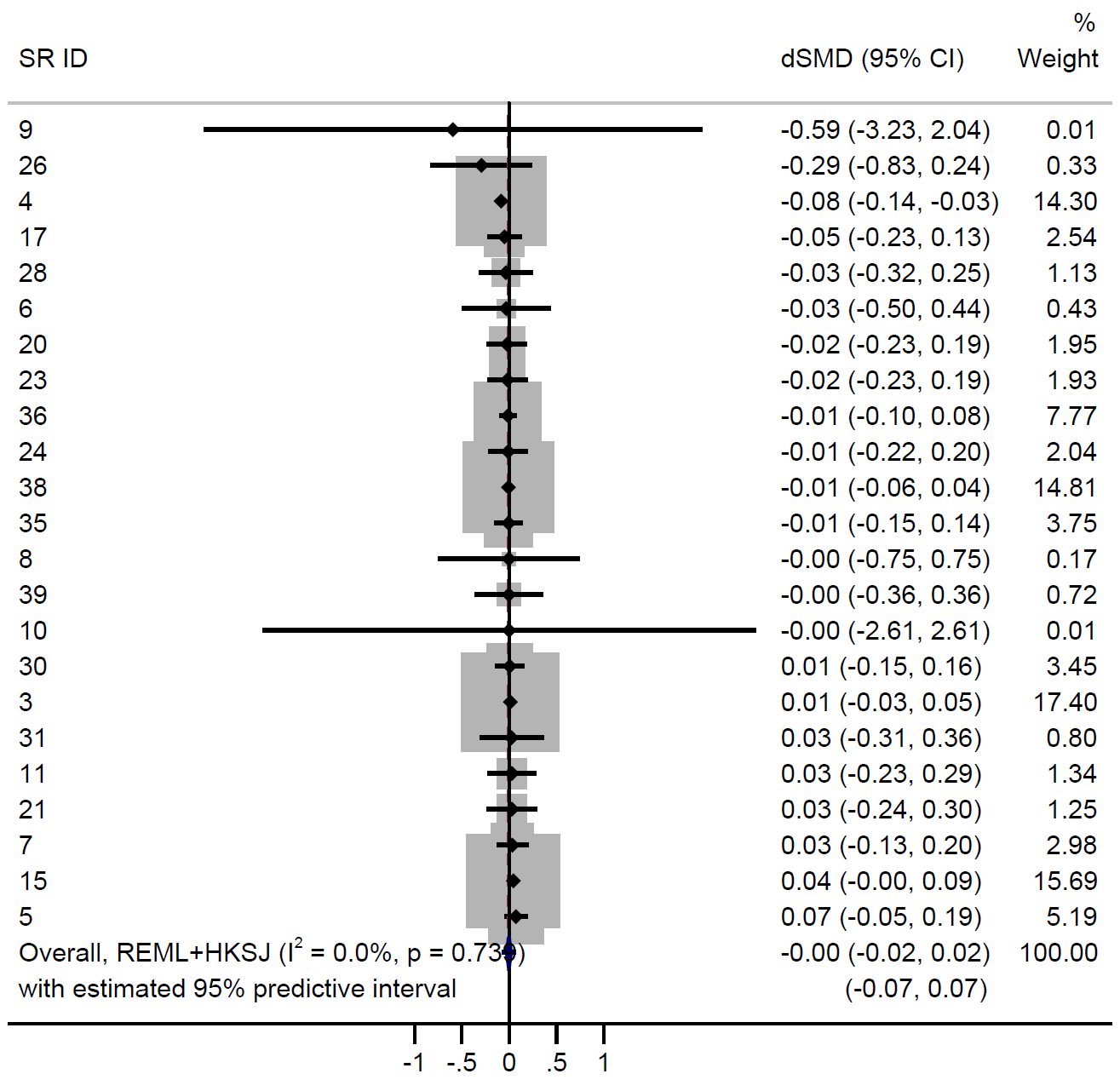

**Supplementary Figure S6. Meta-analysis of differences between the index meta-analytic standardised mean difference (SMD) and median of all its possible meta-analytic SMDs (each calculated using the random-effects model), excluding the meta-analyses in which we assumed risk ratios and hazard ratios provided a good approximation to the odds ratios prior to converting log odds ratios to SMDs. Differences less than zero indicate that the index meta-analytic SMD is more favourable to the intervention compared with the median meta-analytic SMD. dSMD = difference in standardised mean difference.**

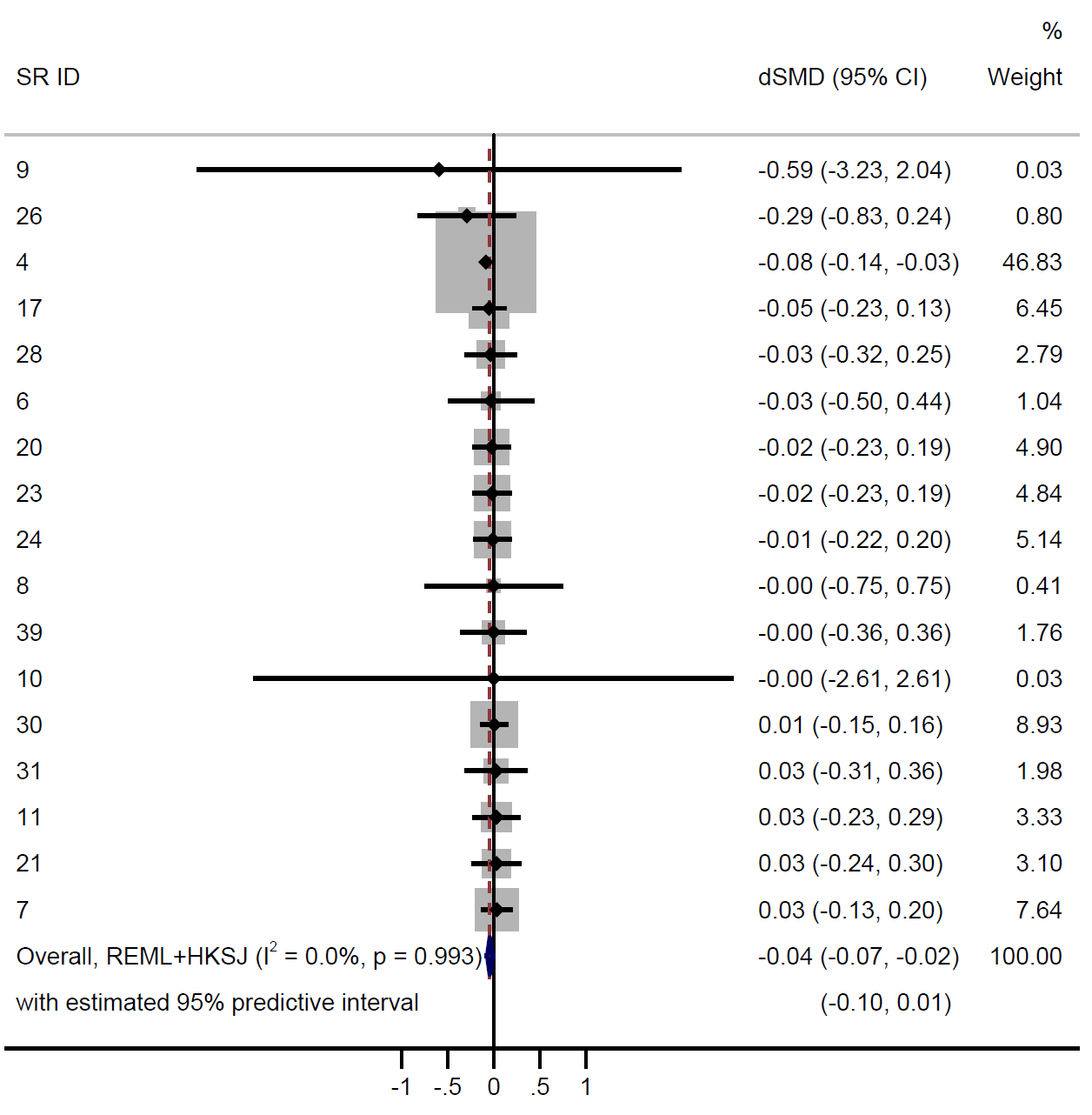

**Supplementary Figure S7. Meta-analysis of differences between the index meta-analytic standardised mean difference (SMD) and median of all its possible meta-analytic SMDs (each calculated using the random-effects model), excluding the meta-analyses of non-continuous outcomes (for which we converted log odds ratios to SMDs). Differences less than zero indicate that the index meta-analytic SMD is more favourable to the intervention compared with the median meta-analytic SMD. dSMD = difference in standardised mean difference.**

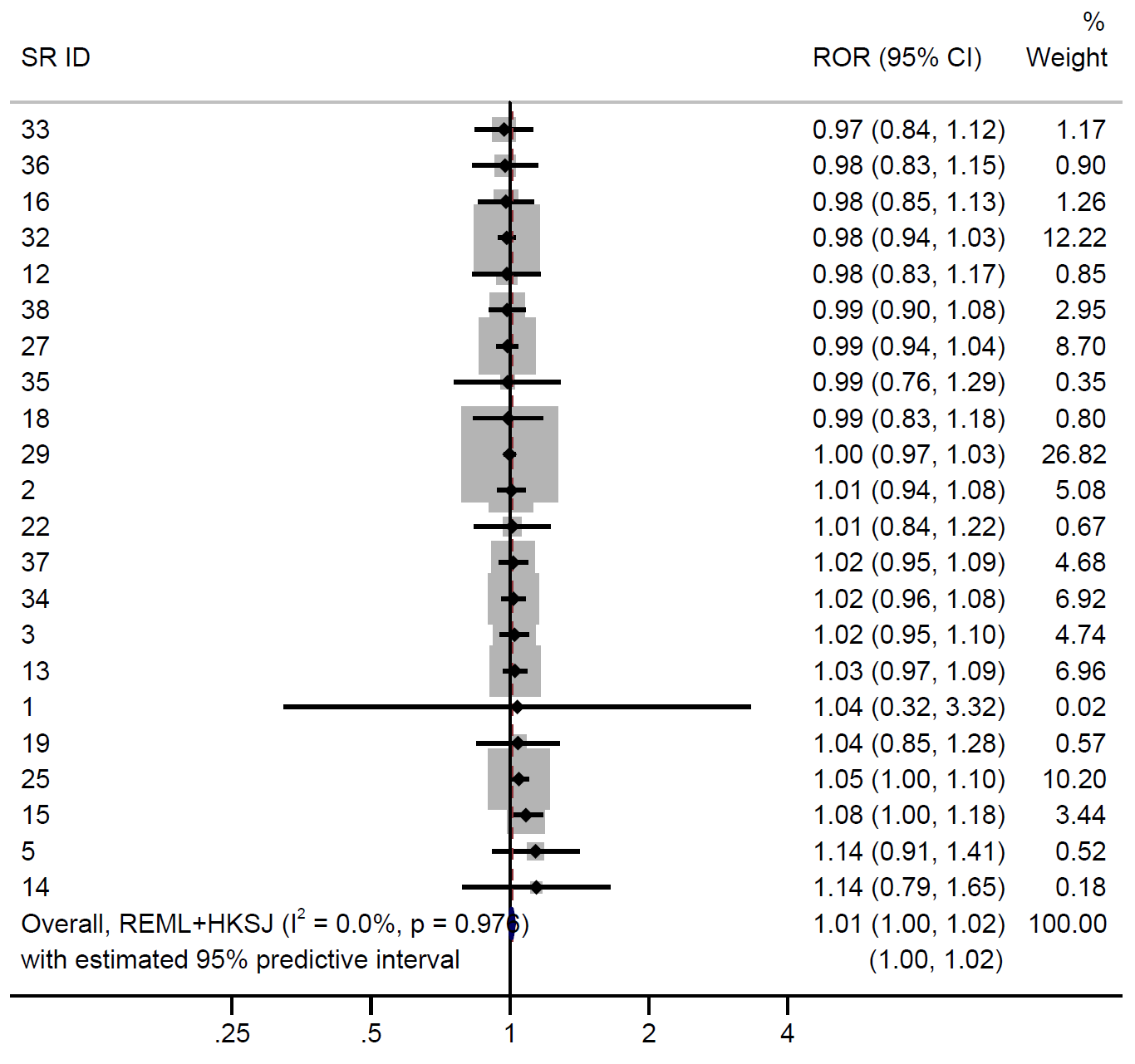

**Supplementary Figure S8. Meta-analysis of differences between the index meta-analytic odds ratio (OR) and median of all its possible meta-analytic ORs (each calculated using the random-effects model), excluding the meta-analyses of continuous outcomes (for which we converted standardised mean difference to log ORs). Differences less than 1 indicate that the index meta-analytic OR is more favourable to the intervention compared with the median meta-analytic OR. ROR = ratio of odds ratios.**

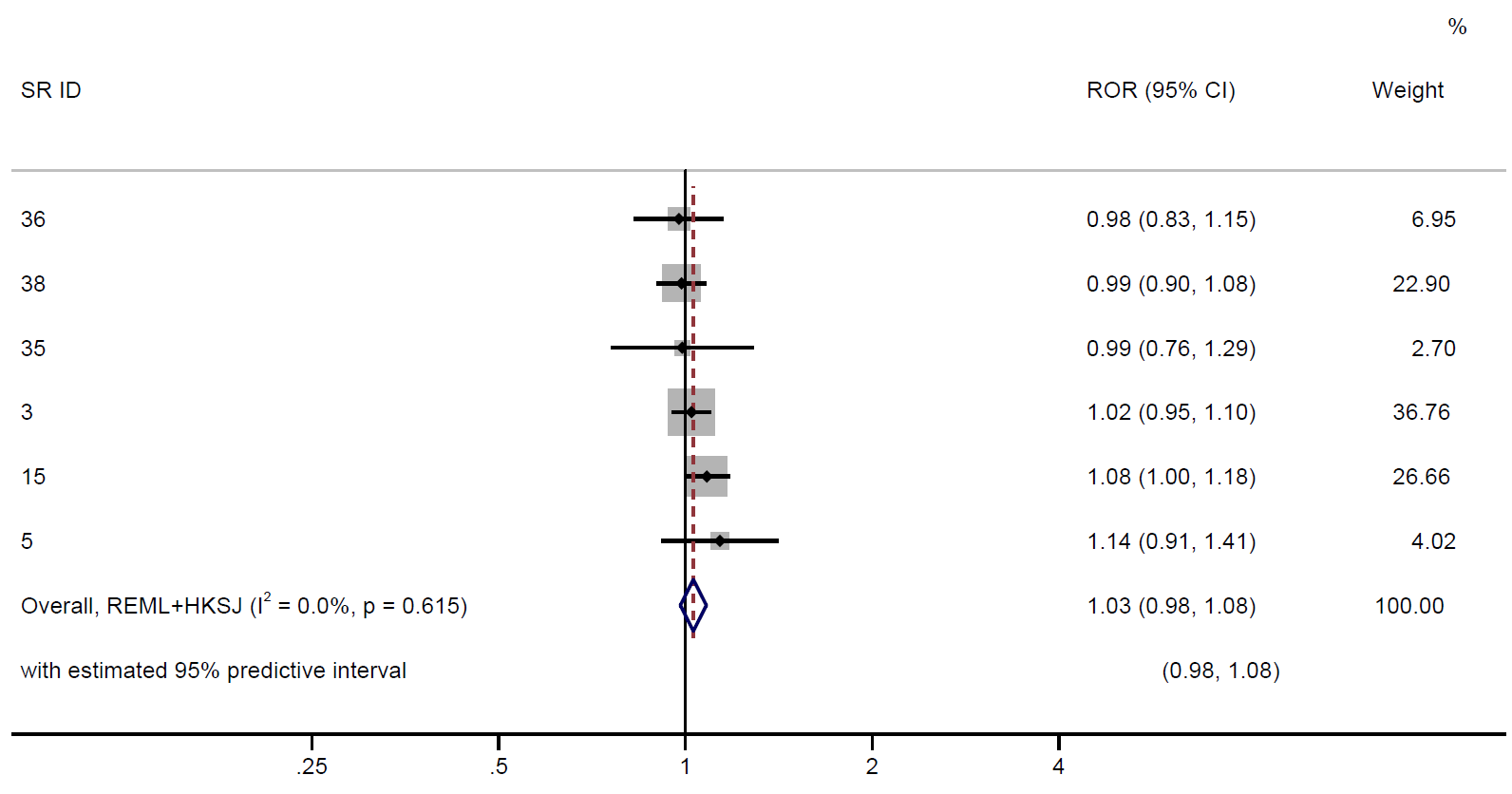

**Supplementary Figure S9. Meta-analysis of differences between the index meta-analytic odds ratio (OR) and median of all its possible meta-analytic ORs (each calculated using the random-effects model), excluding the meta-analyses of continuous outcomes (for which we converted standardised mean differences to log ORs) and meta-analyses in which we assumed risk ratios and hazard ratios provided a good approximation to the odds ratios. Differences less than 1 indicate that the index meta-analytic OR is more favourable to the intervention compared with the median meta-analytic OR. ROR = ratio of odds ratios.**

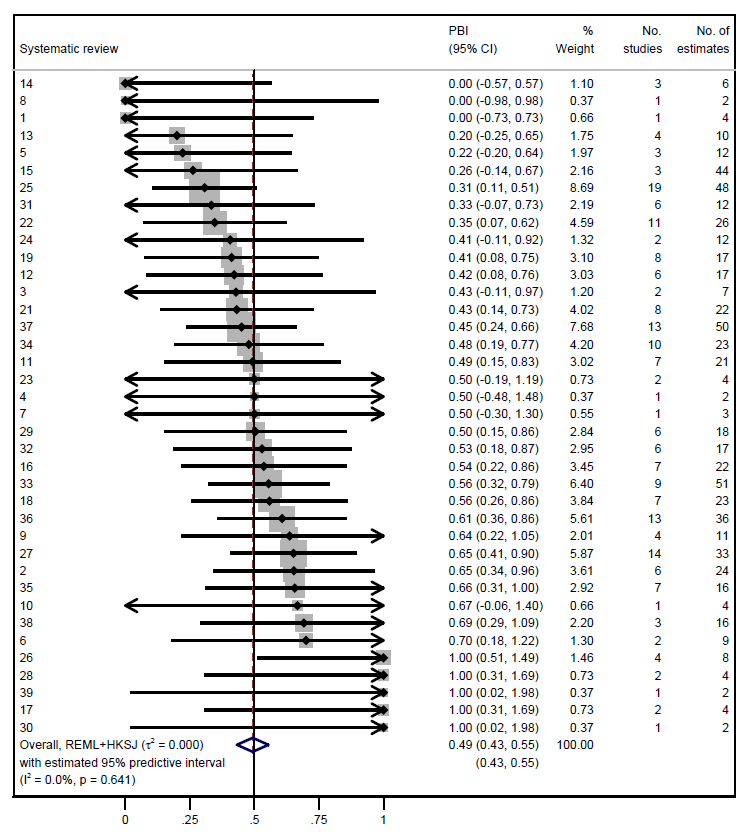

**Supplementary Figure S10. Random-effects meta-analysis of estimates of the PBI per meta-analysis. Systematic reviews are ordered by the magnitude of the PBI estimates.**
